# Multi-omics integration in Esophageal Adenocarcinoma reveals therapeutic targets and EAC-specific regulation of protein abundances

**DOI:** 10.1101/2022.11.24.22281691

**Authors:** J Robert O’Neill, Marcos Yébenes Mayordomo, Goran Mitulović, Sofian Al Shboul, Georges Bedran, Jakub Faktor, Lenka Hernychova, Lukas Uhrik, Maria Gomez-Herranz, Mikołaj Kocikowski, Vicki Save, Bořivoj Vojtěšek, Mark Arends, OCCAMS consortium, Ted Hupp, Javier Alfaro

## Abstract

**Background:** Efforts to address the poor prognosis associated with esophageal adenocarcinoma (EAC) have been hampered by a lack of biomarkers to identify early disease and therapeutic targets. Despite extensive efforts to understand the somatic mutations associated with EAC over the past decade, a gap remains in understanding how the atlas of genomic aberrations in this cancer impacts the proteome. Differences in transcript and the corresponding protein abundances remain under-explored, leaving gaps in our understanding of the mechanisms underlying the disease.

**Methods:** We performed a quantitative proteomic analysis of 23 EACs and matched adjacent normal esophageal and gastric tissues. We explored the correlation of transcript and protein abundance used tissue-matched RNAseq and proteomic data from 7 patients and further integrated these data with a cohort of EAC RNA-seq data (n=264 patients), whole-genome sequencing (n=454 patients) and external published datasets.

**Results:** We quantified protein expression from 5897 genes in EAC and patient-matched normal tissues. Several biomarker candidates with EAC-specific expression were identified including the transmembrane protein GPA33. We further verified the EAC-enriched expression of GPA33 in an external cohort of 115 patients and confirm this as an attractive diagnostic and therapeutic target. To further extend the insights gained from our proteomic data, an integrated analysis of protein and RNA expression in EAC and normal tissues revealed several genes with poorly correlated Protein and RNA abundance, suggesting post-transcriptional regulation of protein expression. These outlier genes including SLC25A30, TAOK2, and AGMAT, only rarely demonstrated somatic mutation suggesting post-transcriptional drivers for this EAC-specific phenotype. AGMAT was demonstrated to be over-expressed at the protein level in EAC compared to adjacent normal tissues with an EAC-specific post-transcriptional mechanism of regulation of protein expression proposed.

**Conclusions:** By quantitative proteomic analysis we have identified GPA33 as an EAC-specific biomarker. Integrated analysis of proteome, transcriptome, and genome in EAC has revealed several genes with tumor-specific post-transcriptional regulation of protein expression which may be an exploitable vulnerability.

## Background

Esophageal cancer is the seventh most common cancer worldwide and the sixth leading cause of cancer death^1^. Esophageal adenocarcinoma (EAC) is now the prevalent histological subtype in western countries and continues to increase in incidence^2, 3^. Gastro-esophageal reflux is thought to be the main driver of the development of EAC as refluxed bile and gastric acid combine to create a carcinogenic environment in the distal esophagus. In this environment, the normally squamous lined esophagus undergoes a metaplastic change to columnar mucosa, known as Barrett’s esophagus, which can progress over time through dysplasia to EAC^4^. If treated by resection at a mucosal-confined, often asymptomatic, early stage the 5-year overall survival approaches 90%^5^. However, despite recent advances in the detection and surveillance of Barrett’s esophagus, the majority of patients with EAC still present with symptomatic, advanced disease, and the prognosis remains grave with a five-year overall survival of less than 20%^6, 7^.

Encouraging advances have been made in systemic chemotherapy regimens for EAC^8^, yet most EACs remain chemotherapy-resistant. Combined with the lack of validated early detection methods, the late presentation of symptomatic cancers and the aggressiveness of this tumor type combine to lead to a highly lethal disease^9^. Identifying an EAC-specific biomarker could improve the detection of invasive disease within a premalignant Barrett’s esophagus segment using either advanced imaging techniques during endoscopy or on ex-vivo cytology specimens or have application in imaging for cancer staging or as a therapeutic target^10^.

Despite knowledge of the genetic changes in EAC developed during large-scale sequencing efforts, no dominant driver oncogene or EAC-specific biomarker has been identified^11^ and this impairs efforts to develop screening tools. Many imaging tools are directed at protein targets yet few studies of global protein abundances have been performed in EAC. Shotgun proteomics is an ideal technique to explore tissue protein abundances in a hypothesis-free context^12^. Our previous work remains one of the most extensive proteomic studies to date in EAC and explored protein abundances by shotgun proteomics in 7 patients^13^. In this current study, we significantly extend this to 23 patients but also improve proteome coverage by leveraging newer-generation mass spectrometry instruments.

Previous studies have explored EAC pathogenesis by genomic or transcriptomic methods^14, 15^, or proteomics in isolation^16, 17^. With developments in sequencing and proteomic techniques, combining DNA, RNA sequencing and proteomic analysis from the same tissue has become a realistic prospect and can illuminate mechanisms of protein abundance regulation and potentially reveal EAC-specific biomarkers obscured in previous single-omic studies.

In this study, we characterize changes in protein and RNA abundance in EAC and generalize these results to a broader cohort. We identified EAC-specific changes in the ratio of protein to RNA abundance which we propose originate through post-transcriptional regulatory mechanisms. These new insights may allow the development of EAC-specific diagnostic or therapeutic tools.

## Methods

### Clinical Samples

Fresh biopsies surplus to diagnostic requirements representing tumor, proximal normal squamous esophagus, and distal normal gastric tissue were collected from resection specimens from patients undergoing surgery for locally advanced esophageal or esophagogastric junctional adenocarcinoma within 30 minutes of specimen resection. Biopsies were snap-frozen in liquid nitrogen and maintained at −80°C until processing. All biopsies were de-identified at the time of collection, subjected to frozen section and histopathological review by a specialist upper gastrointestinal pathologist to confirm tissue of origin and only tumor samples with >50% tumor cellularity proceeded to analysis. In some cases, sufficient paired tissue samples were available for DNA and RNA extraction and sequencing.

### Tissue processing for proteomics

Tissue samples (median mass = 17mg) were processed to tryptic peptides as previously described^13^ (detailed protocol in **Supplementary methods**). Briefly, samples were homogenized using a bead-mill, lysed in 4% SDS lysis buffer at 20:1 buffer to biopsy mass, sonicated, and buffer-exchanged using 30kDa cut-off spin columns (Millipore, MA, USA). Buffer-exchanged lysates were reduced and alkylated before protein concentration determination using the RC-DC^TM^ Protein Assay (BioRad, CA, USA) according to the manufacturer’s instructions. Lysates were digested overnight at 37°C using trypsin (sequencing grade, Promega, Madison, WI) at a 1:100 enzyme to protein ratio (*w/w*). Tryptic peptides solutions were lyophilized using a SpeedVac concentrator (Savant SPD121P, Thermo Scientific, MA, USA) and resuspended in 50 μl 100 mM triethylammonium bicarbonate prior to Tandem Mass Tag (TMT) labeling.

### TMT labeling

TMT label tags were individually resuspended in 41 μl of anhydrous acetonitrile at room temperature (RT). Tryptic peptide samples were labeled with 20.5 μl of the corresponding TMT label reagent (**Supplementary figure 1**) at RT for one hour before reaction quenching using 4 μl of 5% hydroxylamine for 15 minutes. TMT-labeled samples were pooled in low peptide retention tubes (Pierce, Rockford, IL), lyophilized, reconstituted in 0.1% (*v/v*) formic acid in MS-grade water (ThermoFisher Scientific, Loughborough, UK), desalted using Micro SpinColumns C18 (Harvard Apparatus, MA, USA) and lyophilized again.

### Fractionation

The samples from seven initial patients described in our prior published work were fractionated by OFF-GEL electrophoresis as previously described^13^. Subsequent pooled TMT-labeled samples for this work were fractionated using the Pierce^TM^ high-pH Reversed-Phase Peptide Fractionation Kit (Thermo Fisher Scientific, Loughborough, UK) into 8 peptide fractions according to the manufacturer’s instructions (**Supplementary Methods**).

### Mass spectrometry

Individual peptide fractions were subjected to mass spectrometry (MS) using a Q-Exactive Orbitrap Plus equipped with the Flex nano-ESI source and stainless-steel needle (20 µm ID x 10 µm tip ID). The needle voltage was set to 3.1 kV, scan range was 200-2000 m/z. Full MS resolution was set to 70,000, automated gain control (AGC) target to 3×10^6^, and maximum injection time was set to 50ms. For MS/MS analysis, the mass resolution was set to 35,000, the AGC target to 1×10^5^, and the maximum injection time to 120 ms. The isolation width for MS/MS was set to m/z 1.5, and the top 15 ions were selected for fragmentation, single charged ions and ions bearing a charge higher than +7 were excluded from MS/MS. Dynamic exclusion time was set to 20 seconds.

### Database searching and peptide identification

Data have been included in this work from samples from a total of 23 patients with EAC analyzed by TMT LC-MS/MS. Raw peptide reporter ion data were obtained for 7 patients from our previous publication and we generated new data for 16 patients. All subsequent steps in the analysis included all 23 patients. For the 16 patients in this study, ThermoFisher RAW files were converted using msconvert^18^ to mzML files. Database searches were performed using MS-GF+^19^ (v2019.04.18) against a custom database of Ensemble v94 proteins concatenated with reverse decoy sequences and common contaminants. Peptide-spectrum matching was conducted with the following parameters: 10 parts per million precursor tolerance, tryptic cleavage termini with up to 2 missed cleavages, peptide length of 6-40 amino acids, fixed cysteine carbamidomethylation, fixed TMT 6 plex modification (229.1629 Da) on any Lysine or Asparagine terminus, variable modification (−187.152366 Da) on any Lysine residue to account for acetylation rather than TMT modification, and variable Methionine oxidation. Independent false discovery rate (FDR) calculations were made at each stage of identification using Scavager^19, 20^ (v0.1.29) with a threshold of 1% FDR at the PSM peptide and protein level, and two unique peptides required for protein identification.

### Quantitative analysis of proteomes

For quantitative analysis, reporter ion intensities were extracted from mzML files using the pyOpenMS^21^ package with a 0.01 Dalton tolerance. Summary peptide reporter ion intensities were derived from the sum of all corresponding PSM reporter ion intensity values.

To allow integration with the paired RNA expression data and avoid confounding from the uncertainty in transcript and protein isoform identification, all peptides mapping to a unique gene (Ensembl gene identifier; ENSG) were grouped under that gene identifier. To obtain relative expression between patient-matched tissues for a given gene, the ratio of peptide reporter ion intensities was calculated and then the geometric mean of ratios was calculated for each gene as previously reported^13^. Only peptides unique to an ENSG were used for quantification. A summary relative expression value between tumor and normal esophagus (TvE) and tumor and normal gastric tissue (TvG) was calculated for each gene across patients and technical replicates using a meta-analysis approach with a fixed-effect model as previously described^13^. Welch’s modified t-test was used to test the hypothesis that relative protein abundances in the TvE and TvG ratios were not different from the protein abundances of any technical replicates (TvT, EvE, and GvG). The Benjamini-Yekutieli method^22^ was used to correct for multiple hypothesis testing and the false-discovery rate (FDR) - corrected significance threshold was set to P<0.05.

For the correlation of protein and RNA abundance, the geometric mean of all peptide reporter ion intensities for tumor tissues only was calculated for each ENSG for each patient. The meta-analysis with a fixed-effect model approach was then applied to combine patients, weighting the contribution of each patient according to the inverse of the variance of the geometric mean intensity.

### External proteomic data

Raw peptide abundances from normal tissues were obtained from external published MS data^23, 24^. Peptide intensities were processed using the same principles as the esophageal adenocarcinoma samples, pooling all unique peptides that map for each ENSG identifier and deriving a geometric mean for each ENSG.

To account for differing MS instruments, fractionation methods, and possible experimental batch effects, three sequential steps of normalization were applied to the combined dataset of our study and previous publications. The normalization consisted of a sample loading normalization, followed by the trimmed-median of M-values (TMM), and quantile normalization (code available on request).

### Immunohistochemistry (IHC)

Tissue microarrays (TMAs) were constructed as previously reported^13^. Cores on the array included patient-matched normal esophagus, normal stomach, esophageal adenocarcinoma, normal lymph nodes, and lymph node metastases where present. TMA blocks were sectioned at a thickness of 5 μm and placed on positively charged slides (ThermoFisher Scientific, Loughborough, UK) to maximize core adherence. IHC was performed using BOND III autostainer with antibodies to GPA33 (Abcam, ab108938, 1:250 dilution), (Sigma, HPA018858, 1:100 dilution) or IGF2BP1 (Sigma, HPA002037, 1:500 dilution) incubated for 20 min at room temperature and detection using the Leica Bond Polymer Refine Detection kit (DS9800; Leica Biosystems), following the manufacturer’s instructions. Sections representing normal colonic epithelium and normal tonsillar tissue were stained in parallel as positive and negative controls.

Assessment of the IHC intensity staining of TMA cores was performed by two expert histopathologists reaching consensus scores and these samples were graded 0–3 (0 = nil, 1 = weak, 2 = moderate, 3 = strong). Representative IHC images and corresponding intensity scores are shown in **(Supplementary figure 2a).** Cores with significant artifacts (i.e., folded tissue) or loss of tissue material were excluded from the analysis.

### EAC RNA-sequencing data

RNA expression values were derived by RNA sequencing for 264 EAC samples as part of the ICGC ESAD project as previously described^11^. A total of 7 tumors evaluated by shotgun proteomics in this study also had paired RNA-seq data generated. For these 7 patients, unprocessed FASTQ files were mapped to the hg38 reference genome using STAR aligner (v2.6.1). The resulting BAM files were then used to quantify gene expression with RSEM (v1.3.3), obtaining normalized transcript per million (TPM) values for each unique ENSG.

RNA-expression results from a further 257 EAC samples were derived as described in Frankell *et al.*^11^, and expression values in transcripts per million reads were obtained for each unique ENSG quantified for each additional patient.

### External RNA sequencing data

To complement our study of EAC, transcript expression values (obtained as transcript per million values) from normal esophageal samples and a wide range of other normal tissues were incorporated from two external studies - the Wang et al. dataset^23^ and the Genotype-Expression consortium dataset (GTEx consortium) analyses of normal human tissues^25^. Transcript expression from normal tissues was pooled with EAC samples and normalized together at the ENSG level using the trimmed mean of M-values (TMM) and quantile normalization^26^. The GTEx data were downloaded from the GTEx Portal on 12/13/2018.

### EAC Whole-genome sequencing

Whole-genome sequencing data from 454 esophageal adenocarcinomas generated during the International Cancer Genome Consortium Esophageal Adenocarcinoma cancer genome project (ICGC ESAD) were included in the study. Data from 420 tumors were previously published in Frankell *et al*^11^, while data from 34 newly processed samples were included in this study. Raw unprocessed FASTQ files were aligned to the human reference genome (hg38) using bwa (v0.7.17), and duplicate reads were marked using Picard Tools (v2.18.23). Genomic variants were called using the Genome Analysis Toolkit (GATK) best practices Mutect2 (v.4.1.3) and filtered against a germline sample for each patient. Germline variants were also labeled in the same step using data from the 1000 genomes project^27^. The resulting variant files were annotated using VCF2MAF^28^ combined with Ensembl Variant Effect Predictor and then processed using custom scripts in R version 4.1.0.

## Results

### Aberrant expression in the EAC proteins

To identify proteins with enriched or EAC-specific expression, patient-matched samples representing the primary tumor and adjacent normal esophagus and stomach were collected from resection specimens from 23 patients undergoing esophagectomy for EAC (cohort clinical and pathological characteristics in **Table 1a** and **Table 1b**). A total of 5897 gene products were quantified in at least one patient across the three different tissues (1% FDR at the PSM, peptide, and protein levels). Exploring the expression of proteins between adjacent patient-matched tissues revealed relative over-expression of several proteins in EAC compared to surrounding normal tissues (**Figure 2**). Although data were collected using various experimental procedures, no batch effects were observed following normalization and technical replicate ratios clustered together (**Supplementary figure 3**). Entirely EAC-specific protein abundances would not be revealed by our ratiometric expression analysis, however, direct interrogation of the raw data did not reveal any proteins with only unique peptide reporter ion signatures from one tissue and hence entirely tissue-specific expression. Proteins ubiquitously identified across patients were generally from “housekeeping” or cytoskeletal genes and few proteins with tissue-enhanced expression profiles were detected in more than 75% of patients.

**Figure 1:**
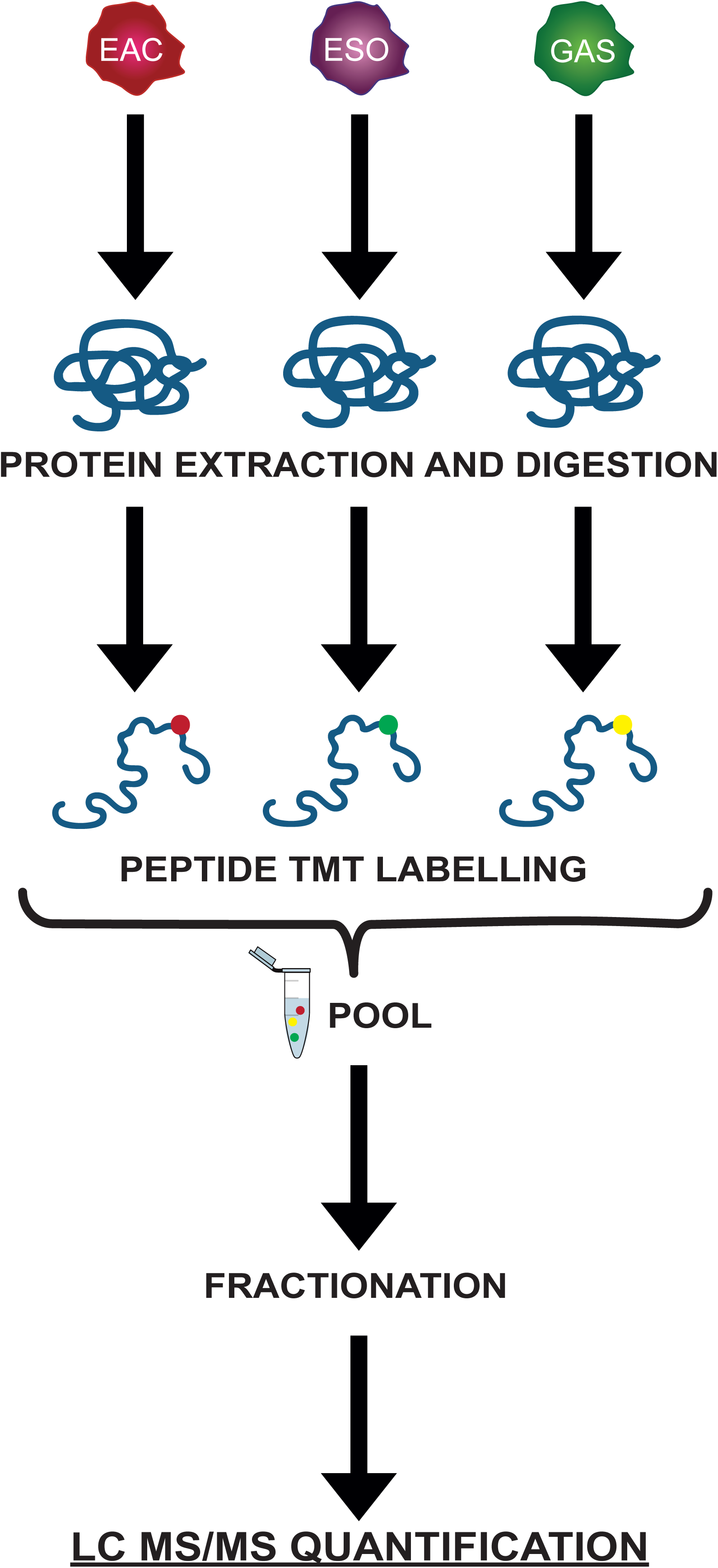
Flow diagram of the proteomic and transcriptomic analysis performed. Global workflow of the proteomic analysis performed using EAC and matched adjacent normal esophagus and normal stomach samples.

**Figure 2.**
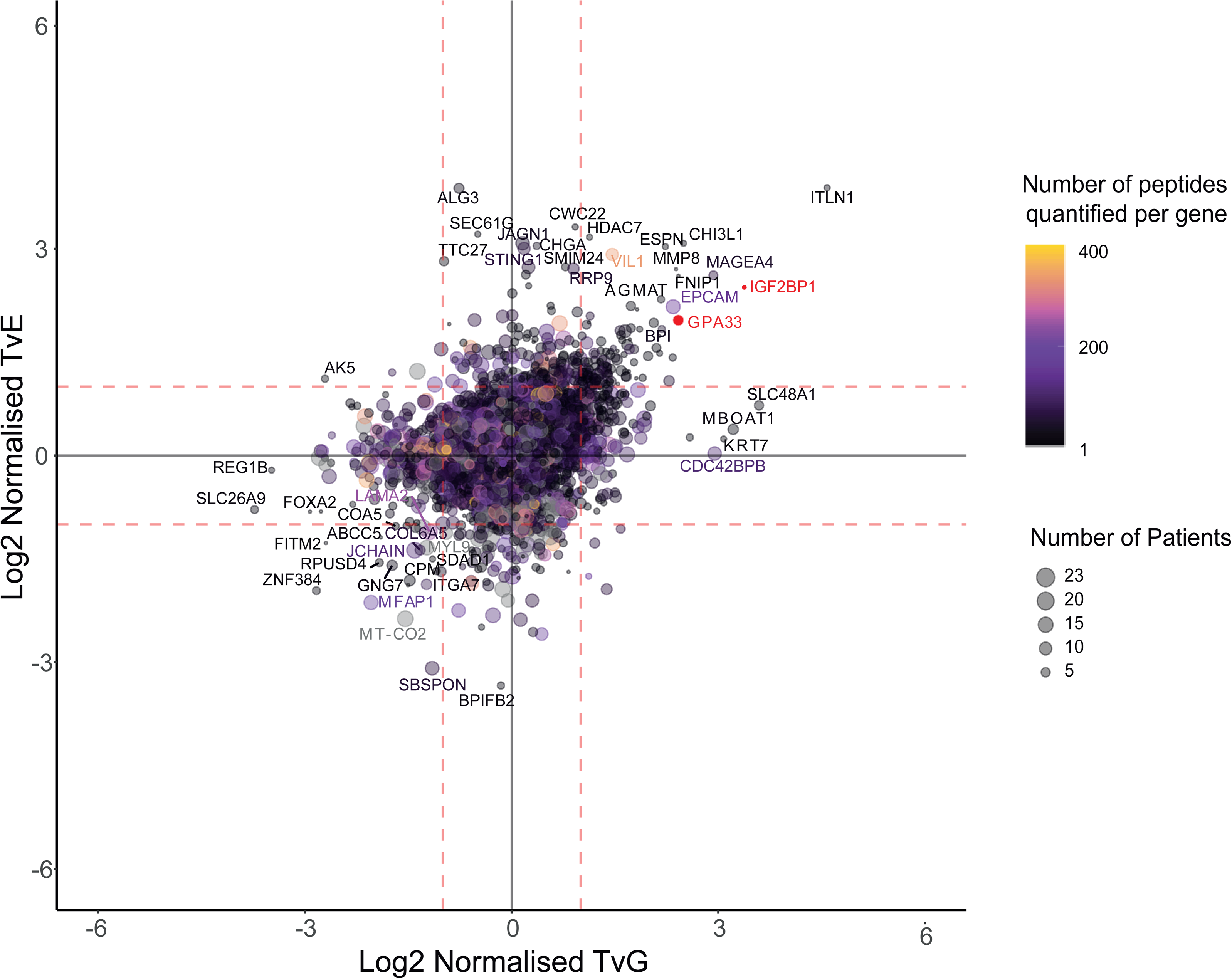
The landscape of protein abundances in EAC relative to patient-matched normal esophageal and normal gastric tissue in 23 patients. Relative expression of 5897 genes across EAC, normal squamous esophagus, and normal stomach in more than one patient. The size of each point indicates the number of patients in which the protein has been quantified and the color represents the number of peptides quantified per protein.

**Table 1:**
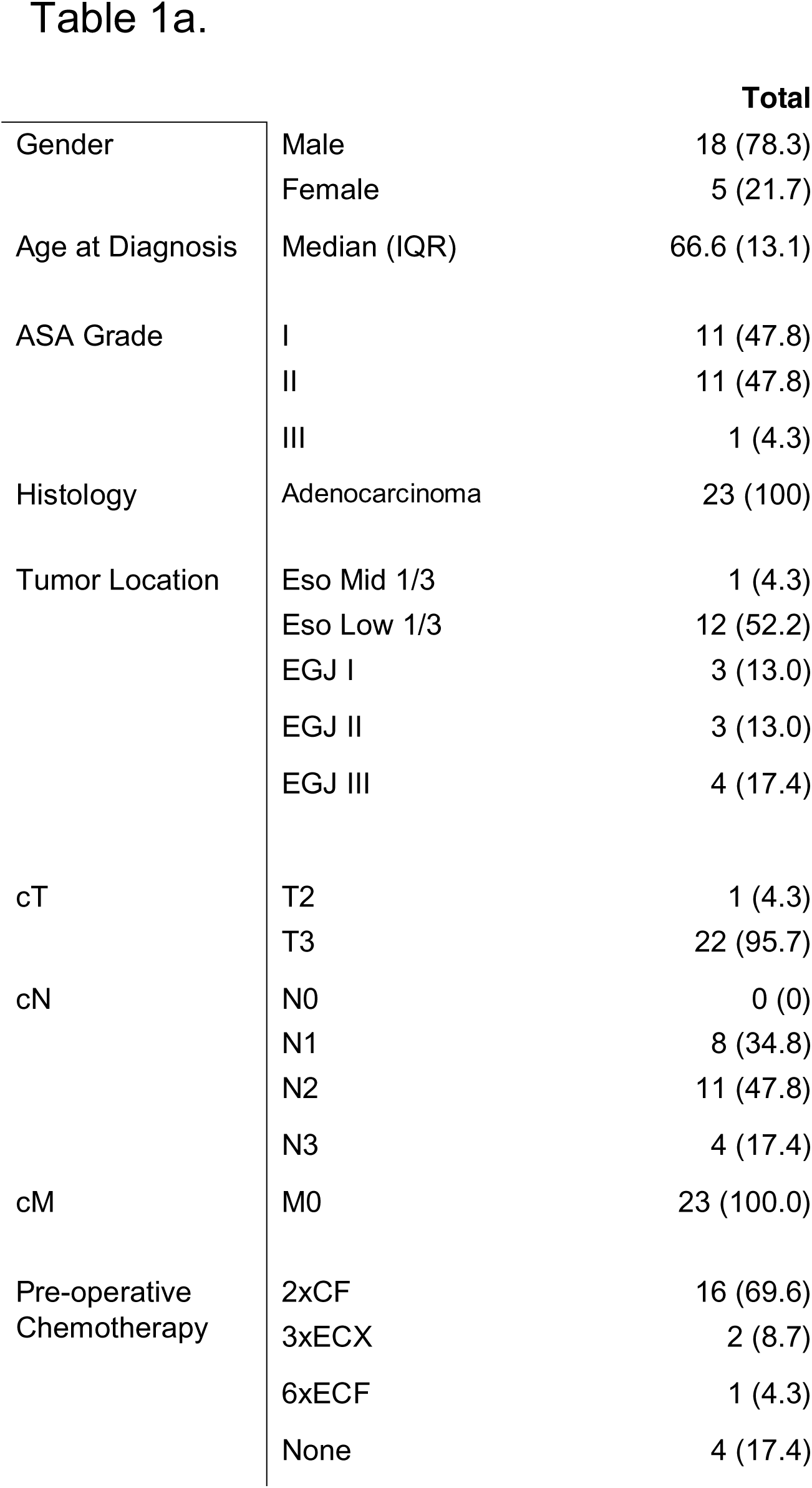
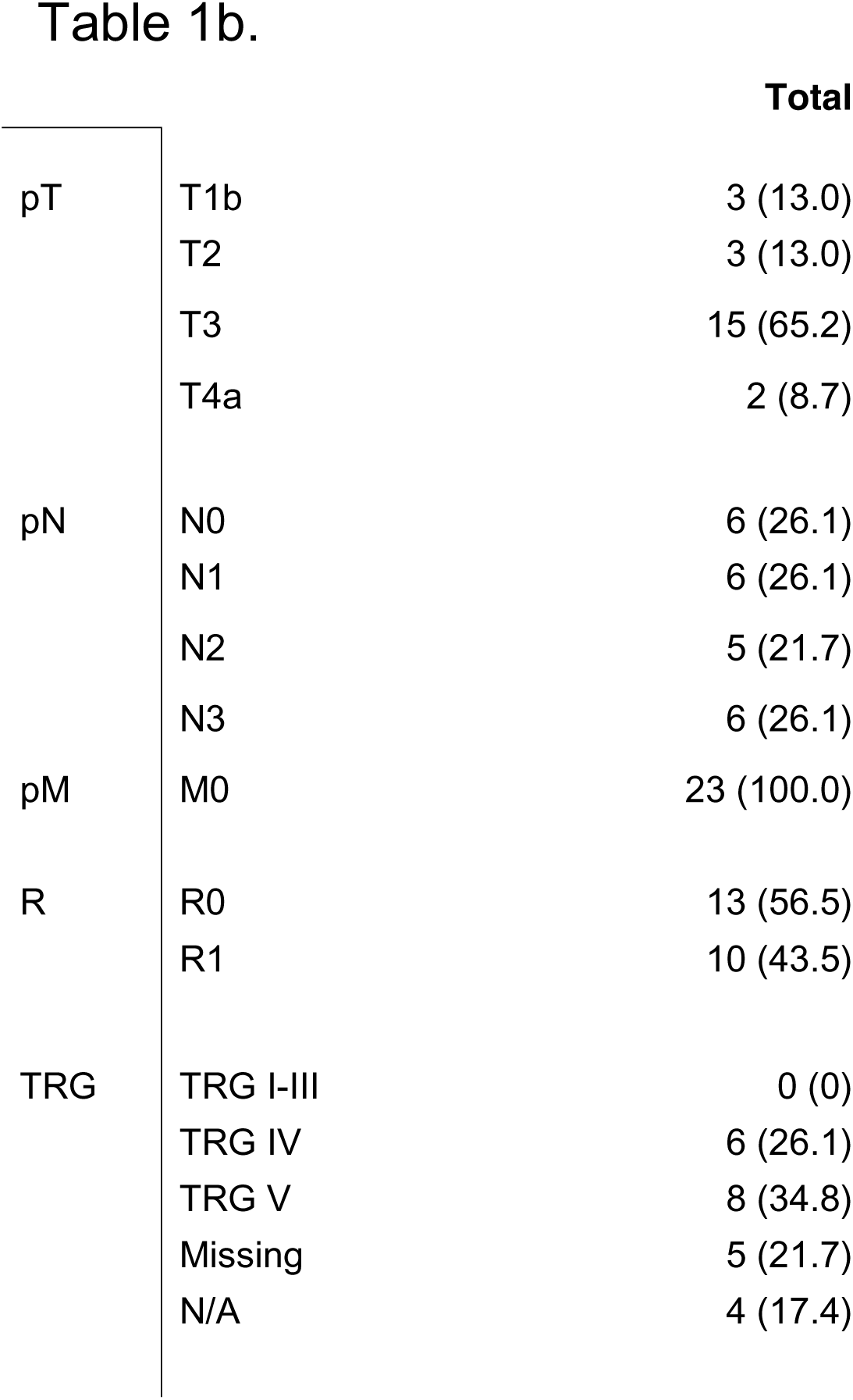
Clinical characteristics of the patients undergoing proteomics analysis. (a) Cohort clinical characteristics. (b) Cohort pathological characteristics.

We once again identified EpCAM as being highly expressed in EAC compared to surrounding normal tissues and have previously validated this expression pattern^13^. Proteins with known expression in intestinal tissues such as Intelectin-1 (ITLN1)^29^ and Villin (VIL1)^30^ were found to be enriched in EAC, likely reflecting the origin of this cancer from intestinal metaplasia of the esophagus^31^. Other proteins previously demonstrated to be over-expressed in EAC were also identified (DSG2^32^, GATA6^11, 33^, or REG4^34^), as well as many other novel genes presenting enriched expression in EAC (**Supplementary table 1**).

We focused on EAC-enriched proteins, defining those as quantified in the majority of our samples (at least 60%) and significantly over-expressed in EAC compared to both normal esophagus and stomach. Nineteen members of the RNA binding motif (RBM) protein family were identified in this study and four (RBM3, RBM6, RBM25, and RBMX) demonstrated EAC-enriched expression. RBM3 was at least 2-fold over-expressed in EAC compared to both normal esophageal and normal gastric tissues.

Cancer-testis antigens have long been held as compelling targets for specific cancer therapies or biomarkers for cancer screening due to their highly restricted tissue expression^35^. We identified several cancer-testis antigens as EAC-enriched including melanoma-associated antigen family members MAGEA4, MAGEA10, MAGED2, and MAGEB2. This group of genes has a well-established role in other cancers and MAGEA4 has been previously demonstrated to be overexpressed in esophageal cancer^36^. Due to its EAC-enriched expression, MAGEA4 is a compelling target for immunotherapeutic approaches. Clinical trials are already underway for MAGEA4-directed adoptive T-cell therapies for patients with MAGEA4-enriched expression in esophageal cancer^37, 38^. For this reason, we evaluated another cancer-testis antigen; IGF2BP1 which was expressed in 10% of patients in this cohort. This gene was over-expressed in EAC compared to patient-matched adjacent normal tissues. There is little published literature on the role of this protein in EAC and we, therefore, sought to validate its expression by immunohistochemistry.

### Validation of GPA33 and IGF2BP1 as EAC-enriched proteins

IGF2BP1 (Insulin-Like Growth Factor-Binding Protein 1) is a single-stranded RNA binding protein with expression in a wide range of fetal tissues and several cancers but the expression in adult tissues is limited to the testis, ovary, prostate, and kidney^39^. A recent study reported higher expression of IGF2BP1 in colonic adenocarcinoma relative to their normal colonic mucosa in a set of 13 paired samples^40^.

The expression of IGF2BP1 in EAC was evaluated by immunohistochemistry using a tissue microarray comprising patient-matched cores from formalin-fixed, paraffin-embedded archival tissues representing primary esophageal adenocarcinoma, lymph node metastases (where present), uninvolved lymph nodes, normal gastric mucosa, and normal squamous esophageal samples. A total of 115 patients’ tissues were included in the array, all of whom had undergone esophagectomy for esophageal adenocarcinoma and 75% had no oncological treatment prior to surgery. The clinicopathological characteristics of these patients have been previously reported^13^.

Across the TMA, IGF2BP1 was highly expressed in 12% of the EAC samples, a similar prevalence to the discovery cohort undergoing shotgun proteomic analysis (identified in 2/23 patients). Expression was absent or very low in normal esophageal squamous tissue **(Figure 3a)**. On analysis of patient-matched cores, there were five EAC cases that showed expression of IGF2BP1 with absent expression in the patient-matched normal squamous tissue **(Figure 3b, C-50, C-25, C-26, C-33, and C-86)**. These observations further support the quantitative proteomic data. High IGF2BP1 protein abundance was identified in 19% of EAC lymph node metastases **(Figure 3a)**. Although around a quarter of uninvolved lymph nodes (5/20 cases) and normal gastric mucosa (7/31 cases) were classified as high IGF2BP1 protein abundances, the staining was confined mainly to a few scattered lymphocytes or potentially non-specific staining within gastric glands (**Supplementary figure 2b**), suggesting that IGF2BP1 tends to be expressed in epithelial-derived tumor cells in EAC. However, more than two-thirds (72%) of the EAC cases showed no protein expression of IGF2BP1 by IHC. Overall, the data suggest that IGF2BP1 protein can be considered to be a moderately tumor-enriched biomarker in EAC but it may only be useful as a therapeutic target or diagnostic biomarker in a limited proportion of patients. The role of IGF2BP1 expression in lymphocytes remains to be determined but may impact the utility of this biomarker in a clinical context for identifying EAC lymph node metastases.

**Figure 3.**
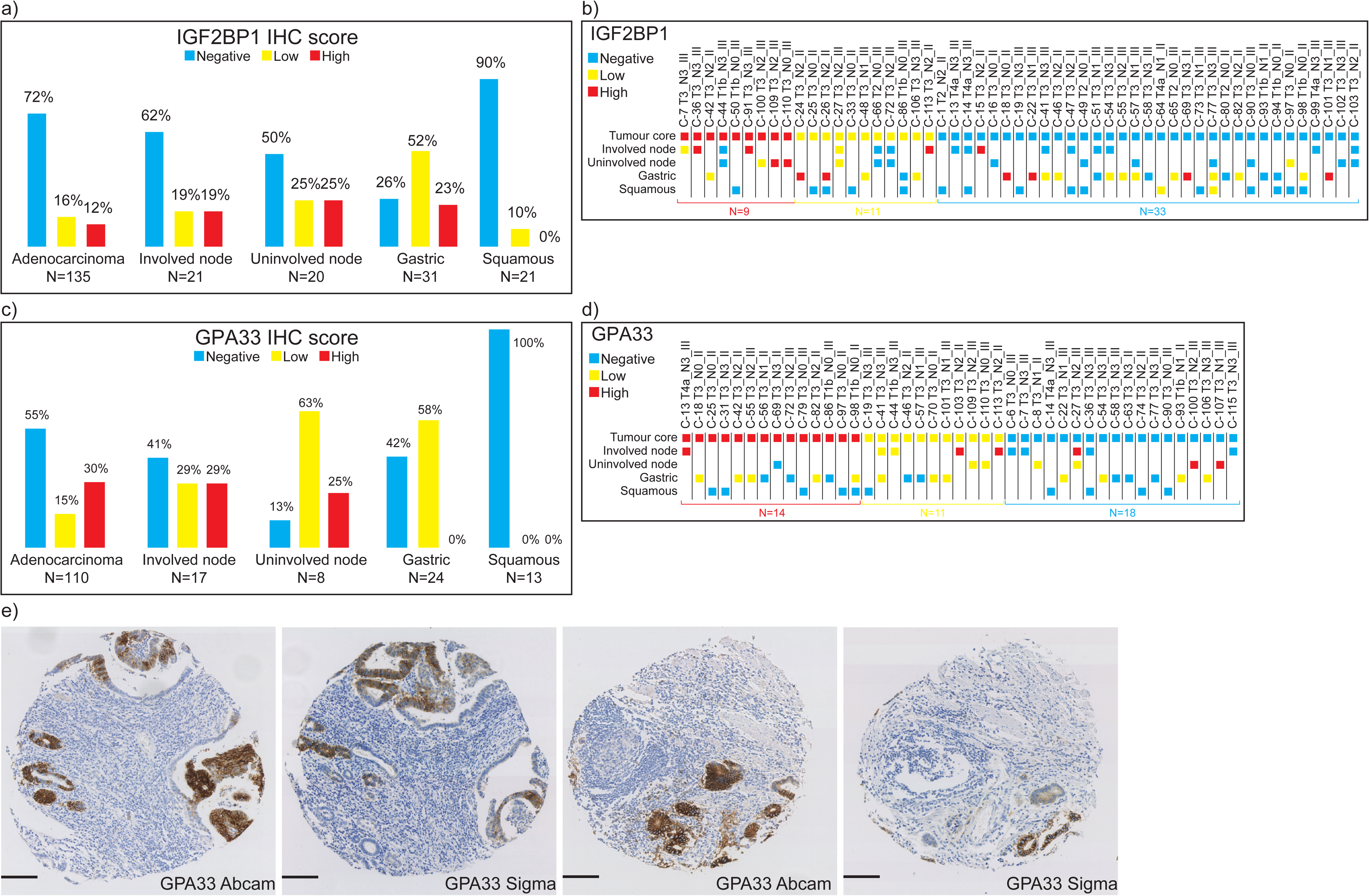
Expression of IGF2BP1 and GPA33 across a tissue microarray containing patient-matched primary esophageal adenocarcinoma, involved (metastatic) lymph nodes, uninvolved lymph nodes, normal gastric, and normal squamous esophageal squamous. **(a)** Immunohistochemistry (IHC) scores for IGF2BP1 protein abundances according to tissue type. **(b)** Scoring according to patient-matched tissue samples including tumor cores and/or involved and uninvolved lymph nodes along with normal gastric and normal esophageal squamous tissues (total n=53). **(c)** IHC scores for GPA33 protein abundances according to tissue type. **(d)** Scoring according to patient-matched tissue samples including tumor cores and/or involved and uninvolved lymph nodes along with normal gastric and normal esophageal squamous tissues (total n=43). **(e)** Representative GPA33 IHC images showing similar staining patterns of two different anti-GPA33 antibodies. IHC staining was evaluated according to DAP intensity. In (b) and (d), each core is identified by a core number followed by pathological T-stage, N-stage, and tumor grade.

Glycoprotein A33 (GPA33) is a cell surface glycoprotein that has been identified as expressed in intestinal tissues and colorectal and gastric cancer and has a putative role in cell adhesion^41, 42^.

GPA33 was highly expressed in around a third of the EAC samples (33/110 cases) in the TMA **(Figure 3c)** which is again consistent with the prevalence in our proteomic cohort (EAC-enriched expression in 8/23 patients). In contrast, only 2 cases across all of the normal tissues (normal lymph node, normal gastric mucosa, and normal esophageal squamous epithelium) showed similarly high GPA33 expression **(Figure 3c Uninvolved node)**. Of the 43 patients with multiple tissue types assessable by IHC, 14 (33%) showed high GPA33 protein levels in primary esophageal adenocarcinoma. There was good concordance with primary and metastatic lymph nodes with either high primary and lymph node expression (1/1 patients, **Figure 3d, C13**) or low primary and either low or high lymph node expression (4/4 patients, **Figure 3d, C41, C44, C103, C113**). For the patients with high GPA33-expressing primary tumors, no normal tissues had high expression. Similarly in all assessed normal tissues (normal lymph node, normal gastric mucosa, and normal esophageal squamous epithelium), there were only 2 cases of high GPA33 staining **(2/37 cores, Figure 3d, C-100, C-107)**. In both of these uninvolved lymph nodes, the staining appeared to arise in a few lymphocytes (**Supplementary figure 4**). We further verified the staining pattern with a second anti-GPA33 antibody **(ab108938, Figure 3e)** on a subset of the cohort (n=62 patients) and found a highly significant correlation in staining scores (Rho=0.822, P<0.001). Overall, these data demonstrate that GPA33 has high EAC-specificity and is worthy of further exploration as an EAC biomarker and potential therapeutic target.

### Correlation of RNA and Protein abundances in EAC

The correlation between RNA abundance and protein abundance has been explored in mammalian cell systems and using global methods in normal human tissues and cancer^23–25, 43^. Uncovering tissue-specific patterns of the regulation of protein abundance independent of RNA abundance implies post-transcriptional regulatory mechanisms which could represent disease biomarkers or drug targets. The control of protein abundance in normal and diseased tissues is still largely unexplored for many genes and if occurring at the post-transcriptional level would limit the relevance of transcriptome analysis to understanding disease biology. Conversely, the greater genome coverage of RNA sequencing provides a distinct advantage over shotgun proteomics and this technique may be better suited to future translational studies in the case that protein and RNA abundance are well correlated. To explore this in EAC, we correlated the expression of protein and mRNA in seven patients in our proteomic discovery cohort who had sufficient tumor tissue to undergo both RNA sequencing and shotgun proteomics (**Supplementary figure 5a-h**). These tissue-matched datasets present a unique opportunity to explore the decoupling of RNA and protein abundance.

We quantified both RNA and protein abundance in EAC tissues in at least one patient across 5,531 genes. Although genome coverage and the strength of correlation varied across patients there was a moderate correlation between RNA and protein abundance overall (Pearson’s value of 0.46), in keeping with previous reports from other human tissues^23^.

### Evaluation of Protein to RNA expression ratios in EAC tissue

By plotting genes according to both protein and RNA expression we identify a group of 15 outlier genes with low protein to RNA expression ratios (Log_2_ Protein intensity <26, Log_2_ RNA TPM >10; **Figure 4a, highlighted blue)**. Although these genes may have been identified in a transcriptome analysis as highly expressed, they are less likely to be of interest from a biomarker perspective due to low relative protein expression. In contrast, we also identified a group of 30 outlier genes with high protein-to-RNA expression ratios (Log_2_ Protein intensity >28, Log_2_ RNA TPM <8; **Figure 4a, highlighted red**). These may not have been identified from a transcriptome-only analysis and may well be biomarker candidates or therapeutic targets.

**Figure 4.**
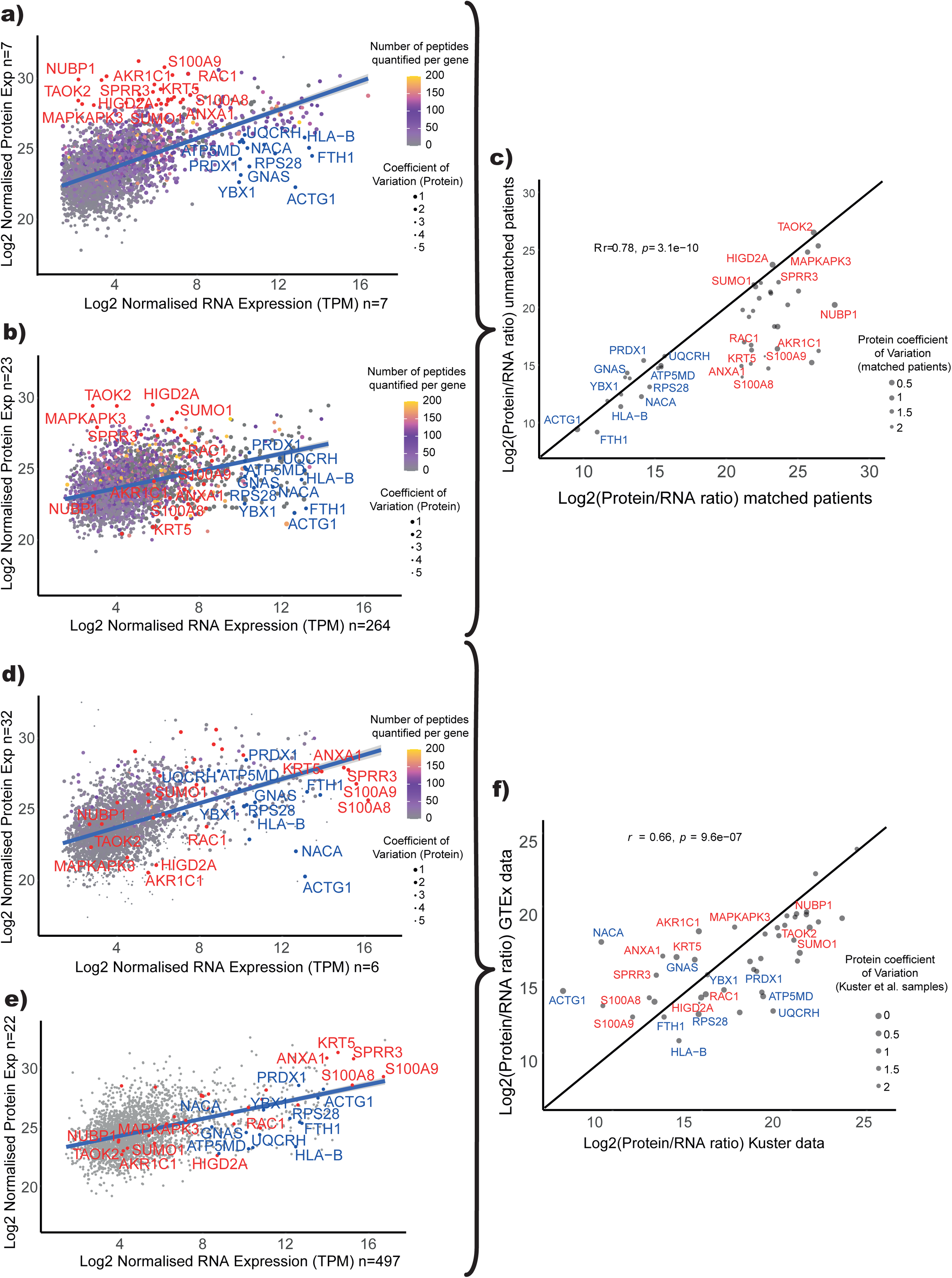
Direct correlation of protein and RNA expression in EAC and normal squamous esophagus. **(a)** Patient-matched expression of RNA and Protein in EAC (n=7 patients). **(b)** Correlation of RNA and protein abundances in unmatched EAC samples (n=23 proteomic data, n=264 transcriptomic data) **(c)** Correlation of outlier protein to RNA ratios in matched and unmatched EAC cohorts. **(d)** Correlation of RNA and protein abundances in the normal squamous esophagus using Wang et al. data. Outliers from (a) are highlighted. **(e)** Correlation of RNA and protein abundances in the normal squamous esophagus using GTEx consortium data. Outliers from (a) are highlighted. **(f)** Correlation of outlier protein to RNA ratios in Wang et al. and GTEx datasets.

Several of these genes with high protein to RNA ratios are of interest including RHNO1, CHFR, and CENPE (**Supplementary figure 5h**). RHNO1 has been recently reported as a prognostic marker in colorectal cancer^44^, renal, and liver cancer, where the high expression of the gene ends in an unfavorable prognosis for the patient^45^. CHFR is an E3-ubiquitin ligase with a role in cell cycle checkpoints and is known to be downregulated EAC at the RNA level^46, 47^. This was again demonstrated in this analysis yet CHFR was highly expressed at the protein level, highlighting the need to corroborate RNA expression results with data at the protein level. Another gene following this pattern of expression is CENPE, which is associated with lung adenocarcinoma cell proliferation^48^, and expression is correlated with poor prognosis in EAC^49^. It is possible that protein abundance is not transcriptionally controlled for these genes and therefore their expression must be studied directly at the protein level.

### Tissue-matched protein to RNA expression ratios are representative of global EAC protein to RNA expression ratios

We have directly assessed protein to RNA expression ratios by undertaking both proteomic and RNAseq analysis from the same tumors from seven patients yet it is not clear if this small sample is representative of the pattern of protein to RNA expression ratio in EAC in general. To assess this we used RNA expression data derived from RNAseq analysis of 264 EACs from patients in the OCCAMS study taking part in the ICGC ESAD project. Protein expression in EAC alone derived from the pooled analysis of the 23 patients presented in **Figure 2** were plotted against RNA expression derived from the OCCAMS cohort on a gene-wise basis (**Figure 4b**).

Most outlier genes with either a high or low protein to RNA ratio identified on the matched tissue analysis presented a consistent ratio in the unmatched analysis (**Figure 4c**, Pearson’s r=0.78, P<0.001). Where differences were observed, this was usually down to a difference in the quantified protein abundances level accompanied by a high coefficient of variation between samples, e.g. NUBP1 or AKR1C1. This may reflect challenges associated with protein quantitation using a shotgun proteomic strategy and our stringent approach using only uniquely identified peptides for protein quantification. We, therefore, conclude that for most of our outlier genes, the protein to RNA ratios identified in EAC are robust.

### Exploring tissue differences in protein to RNA expression ratio

Previous reports from analysis across a range of normal human tissues suggest that protein to RNA ratio is both a surrogate of translation rate and is gene intrinsic. It has been proposed that protein to RNA abundance ratio, although widely variable between genes, is preserved for a specific gene across multiple tissue types^50–52^. We used two external datasets of tissue-matched RNA and proteomic analysis, the GTEx consortium dataset and the Wang et al. dataset to evaluate the protein to RNA ratio of outlier genes identified in our analysis of EAC in normal squamous esophageal tissue.

Outlier genes with consistent protein to RNA ratios across both matched and unmatched EAC cohorts demonstrated significantly different protein to RNA ratios in normal squamous esophageal tissue in both the Wang et al. data (**Figure 4d**) and the GTEx data (**Figure 4e**). Despite disparate experimental strategies and quantitative proteomic methods between these 2 large-scale studies, the protein to RNA ratios for outlier genes were well correlated between studies (**Figure 4f**, Pearson’s r = 0.66, P<0.001). This raises the possibility that there are EAC-specific mechanisms of post-transcriptional control of protein abundance at least for these outlier genes.

Several outlier genes (KRT5, ANXA1, SPRR3, S100A8, and S100A9) demonstrated lower RNA expression in EAC when compared to normal esophageal tissue while still being highly expressed at the protein level in both tissues. RHNO1 was also demonstrated to have very low RNA expression in EAC leading to very high protein to RNA ratios in comparison with normal tissues.

In contrast, when comparing the EAC data with the external normal squamous esophageal data, other dysregulated genes, including TAOK2, MAPKAPK3, and HIGD2A, appear to have a higher protein abundance in EAC compared to normal squamous esophageal tissue while presenting similar transcript expression levels.

### Exploring changes of Protein to RNA expressions across tissues

We next compared protein to RNA ratios for outlier genes in matched and unmatched EAC cohorts and the wider range of normal tissues assessed in the GTEx and Wang et al. studies to determine if these findings were generalizable to other normal human tissues. The protein to RNA ratios across EAC and multiple tissues for several outlier genes are summarised in **Figure 5**. To further explore the potential deregulation of protein from RNA abundance during the development of EAC from Barrett’s, genes associated with intestinal differentiation or Barrett’s esophagus (AGMAT, HMGA1, EPHB2, OLFM4)^31^ were appended to the outliers identified in our matched tissue analysis of EAC.

**Figure 5.**
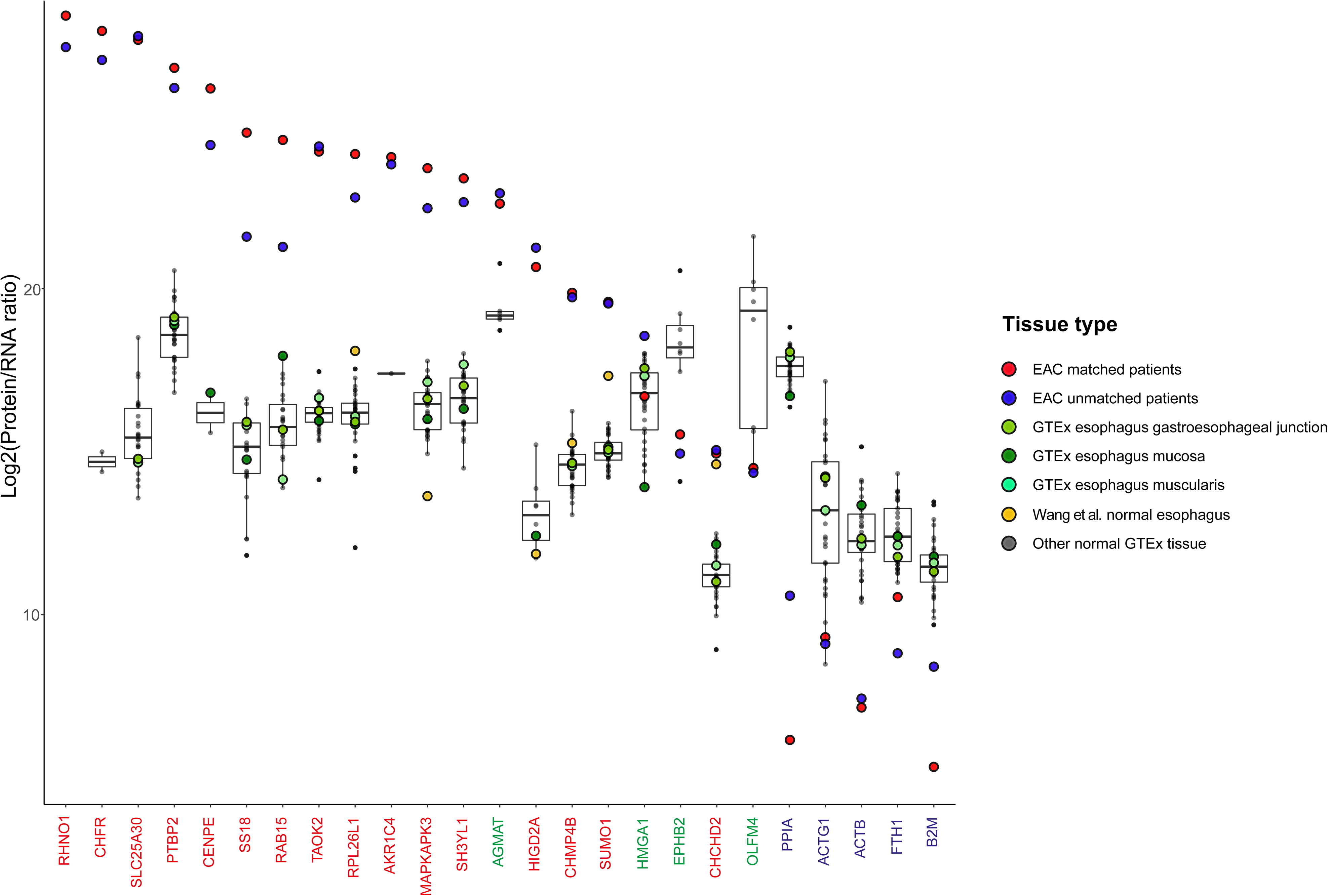
Distribution of protein to RNA ratios for outlier genes across normal tissues and EAC. Each gene represents an outlier gene defined in Figure 4a. Each point represents a tissue type and normal tissue Protein to RNA ratios have been summarized as a boxplot with median, box limits are 25th and 75th centile, and tails represent maximum and minimums values as well as outliers from all normal tissues limits. Dysregulated genes with high protein abundances and low RNA expression were colored in red, as well as genes associated with intestinal differentiation or Barrett’s esophagus (green) and genes with low protein abundances and high RNA expression (blue).

The striking finding is that the protein to RNA ratio is similar for each gene across a wide range of normal tissues, in keeping with prior reports that this is a gene-intrinsic property. In contrast, however, in our analysis of EAC, there are widely different protein to RNA ratios that are consistent between matched and unmatched cohorts for several outlier genes suggesting this property is not gene-intrinsic for these genes in EAC.

For the genes associated with intestinal differentiation or Barrett’s esophagus (HMGA1, EPHB2, and OLFM4) the protein to RNA ratios were similar between normal tissues and EAC suggesting regulation of protein abundance is constant for these genes during the development of EAC from Barrett’s esophagus. AGMAT, however, has a higher protein to RNA ratio in EAC compared to a wide range of normal tissues suggesting this regulation may be altered during Barrett’s carcinogenesis. We previously demonstrated that AGMAT was significantly over-expressed at the protein level in EAC compared to the patient-matched normal squamous esophagus and gastric tissue (**Figure 2**) and AGMAT has recently been described with a role in cancer by promoting tumorigenesis via MAPK signalling^53^. Together these data confirm AGMAT is over-expressed in EAC, and we propose this over-expression is post-transcriptionally controlled. Further work is required to understand the mechanism and the role of AGMAT in EAC development.

### Dysregulated protein to RNA ratio is not driven by somatic mutation

EAC is associated with a significant tumor mutational burden^54^ and so we next sought to determine if the somatic mutation was correlated with post-transcriptional changes in protein abundance for the outlier genes. WGS data were acquired from EAC tissue from 454 patients sequenced as part of the OCCAMS ICGC ESAD Cancer Genome Project. Tumors were sequenced at a mean depth of 70x coverage in EAC (minimum depth 52x) and a mean depth of 42x coverage in patient-matched germline tissue (minimum depth 29x). Somatic variants were only detected in 15 of the 45 outlier genes and variant allele frequencies ranged from <1 - 3% (**Supplementary figure 6**). This low rate of somatic mutation, despite high tumor sequencing coverage, suggests somatic variants are not the driver of the protein to RNA expression ratio changes for these outlier genes. We, therefore, conclude the EAC-specific post-transcriptional change in protein abundance revealed for the outlier genes is driven by an alternative mechanism.

## Discussion

In this study we describe the expressed protein landscape in EAC and matched adjacent normal tissues from 23 patients undergoing surgical resection for locally advanced disease and describe the relative expression of at least 5897 proteins in these tissues. To our knowledge, this represents the largest characterization of relative protein abundances in EAC in the literature to date. In keeping with other published shotgun proteomic studies, the majority of proteins were only identified in a subset of the 23 patients. This may reflect a combination of low proteome coverage, an intrinsic limitation in the shotgun proteomic strategy, and inter-patient heterogeneity. Despite this, we gain valuable new insights into the expressed protein landscape in this cancer.

We once again confirm EPCAM as highly expressed relative to surrounding normal tissues and have previously validated the EAC-specific expression of EPCAM^13^. We also identified several RBM protein family members as demonstrating EAC-enriched expression including RBM3. In support of this finding, RBM3 has been previously implicated in cancer and reported to be over-expressed in esophageal and gastric adenocarcinoma with particularly high expression in those cancers with a background of intestinal metaplasia^55, 56^.

We report several further proteins with EAC-enriched expression, some of which are novel but others (INTL1, VIL1, OLFM4, REG4, ANXA13) have been previously described as expressed selectively in intestinal tissue or goblet cells, a histological marker of intestinal differentiation. We propose this may reflect the origin of EAC from glandular metaplasia of the esophagus. We further identify and validate the EAC-enriched expression of GPA33 in a third of patients with EAC and the over-expression of IGF2BP1 in 10% of patients with EAC. The limited expression of GPA33 in patient-matched normal tissues provides a compelling basis for further development of this as an EAC biomarker for imaging and cancer detection or development as a therapeutic target. GPA33 has previously been demonstrated to be highly expressed in gastric and colorectal cancer and human anti-GPA33 antibodies are being evaluated as immuno-oncological treatments^57–59^. Early phase clinical trials of monoclonal anti-GPA33 showed good safety and tolerability^60^, while other clinical trials investigating novel anti-GPA33 antibodies in colorectal cancer are ongoing (NCT02248805). Although we did not identify any background normal squamous expression and only limited non-specific gastric mucosal expression of GPA33, two cores from normal lymph nodes across our validation TMA cohort demonstrated some scattered GPA33-expressing cells, presumed to be lymphocytes (**Supplementary figure 4**). A recent study has confirmed GPA33 expression in a subset of CD4-positive T-regulatory cells with an immunosuppressive phenotype which would support our findings^61^. The role of these cells in cancer remains to be determined.

We further extended our analysis to seven patients in this cohort by quantifying RNA expression by RNAseq in the same EAC tissues subjected to quantitative shotgun proteomics. We initially explored the correlation between protein and RNA abundance in this tissue. The changes in expression between protein and RNA abundances and how they are regulated as part of normal tissue homeostasis are still being delineated^62^, and are only recently being evaluated in malignancy^63^. However, it is clear that both protein degradation and synthesis pathways can be tightly regulated^64^. The former through post-translational modifications of proteins targeting them towards proteasomal degradation and the latter arises through post-transcriptional regulation of the epitranscriptome decorrelating RNA and protein abundances. Mutation, copy number variation, and epigenetic modification are well established to modify protein abundances, and regulation of protein abundance via post-translational or post-transcriptional pathways is likely another exploitable mechanism.

The combined analysis of protein and RNA expression in tissue-matched EAC patients revealed two groups of outlier genes with either high or low protein to RNA expression ratios. We validated these findings by comparison of protein abundance in EAC from our full proteomic cohort of 23 patients with RNA abundance from 264 EACs. We demonstrated an excellent correlation in protein to RNA ratios between the matched and unmatched cohorts for these outlier genes suggesting our cohort of seven patients was a representative sample of EAC.

We next sought to determine if the protein to RNA ratios observed for the outlier genes in EAC were consistent across other tissue types, and therefore a gene-intrinsic phenomenon, or if these findings were solely observable in EAC; a tissue-specific phenomenon.

There have been two significant efforts to study the correlation of RNA and protein abundances in normal tissue: the GTEx project^24, 25^ and the Wang et al. dataset^23^. We used the available raw data from these studies to evaluate protein to RNA ratios across normal tissues for our outlier genes and found protein to RNA ratios to be well correlated between these studies. Protein to RNA ratios for the EAC outlier genes were similar across a wide range of normal human tissues in support of the previous reports that protein to RNA ratio is gene intrinsic.

To our surprise, several outlier genes had very consistent protein-to RNA ratios across both matched and unmatched EAC cohorts but demonstrated significantly different protein to RNA ratios across all the normal tissues assessed in GTEx and Wang et al. studies. We conclude that, for these genes, protein abundance is controlled by a post-transcriptional mechanism which is altered in EAC. This offers a potentially cancer-specific targetable mechanism and is worthy of further exploration.

Outlier genes were associated with several pathways including those involved in mitochondrial function and the p38/MAPK signaling cascade. Both SLC25A30 and PTBP2 are described to influence mitochondrial function, and promote cancer cell proliferation^65,66^. Prior work has suggested S100A9 is down-regulated in the progression from Barrett’s metaplasia to EAC but this only assessed expression at the mRNA level^67^, in contrast in a further study the protein has been found to be over-expressed in EAC in comparison to Barrett’s tissue samples and upregulated in serum in patients with EAC^68^. Our findings further support these data and emphasize the importance of evaluating protein abundances directly.

Both MAPKAPK3 and TAOK are members of the p38 MAPK signaling pathway which has a well-established role in oncogenesis and is a potential therapeutic target^69, 70^. HIGD2A is upregulated in response to hypoxia^71^, a likely feature of the tumor microenvironment. It is possible for these outlier genes, post-transcriptional regulation of protein abundance differs in EAC. It is not clear if these effects are unique to EAC or more broadly represent a property of malignant or diseased tissue. For selected candidates, altered post-transcriptional regulation may offer a tumor-specific property that could be exploited for diagnostic tools or for therapy. AGMAT was demonstrated to have a high protein to RNA ratio in EAC and a significantly lower protein to RNA ratio in other normal tissues. AGMAT was also over-expressed at the protein level in EAC compared to both patient-matched normal squamous esophagus and normal stomach. Based on these data we propose that the mechanisms of over-expression in EAC is post-transcriptional. The role of AGMAT in EAC and the exact mechanism leading to protein over-expression are worthy of further exploration in follow-up studies.

We have directly assessed global protein abundances in EAC and patient-matched adjacent normal tissues in 23 patients but have only directly assessed RNA expression in the same EAC tissue from seven of these patients. It is possible our findings regarding protein to RNA expression ratio in EAC were limited by statistical power. For this reason, we evaluated RNA expression in a large external cohort of 264 EACs and compared it with protein abundances from our cohort of 23 patients. In support of our findings, there was an excellent correlation in protein to RNA expression between patient-matched and unmatched cohorts.

We have also iterated our proteomic methods across time in collecting these data. From our previous proteomic study, the largest source of biological variation was found to be inter-patient heterogeneity rather than technical variation. To address this we used data from the maximum number of patients rather than selecting those with a more homogenous experimental strategy. In support of our quantitative proteomic findings - many of our EAC-enriched proteins have been previously published as upregulated in EAC and both GPA33 and IGF2BP1 were over-expressed in a very similar proportion of our discovery cohort and the external TMA validation cohort.

The use of diverse proteomic and transcriptomic methods in both GTEx and Wang et al. studies has the potential to confound our conclusions. However, we have applied robust normalization to deal with batch effects and we have only explored relative abundances to further limit the impact of variable quantitative dynamic ranges. A good correlation was achieved for these two studies when comparing outlier protein to RNA ratios in normal squamous esophageal tissue in support of these methods. The use of these large external datasets provided the capacity to explore protein to RNA in a wide range of tissues beyond that achievable using our dataset. We caution that detailed validation of our findings is still required for candidate genes where we propose EAC-specific post-transcriptional regulation of protein abundances. If confirmed, however, this novel finding may provide a tumor-specific targetable mechanism with potential applications in diagnosis and therapy.

## Conclusions

In this study we use proteomic methods to present the landscape of protein abundances in EAC and matched-adjacent tissues. We identify several EAC-enriched proteins and externally validate GPA33 as over-expressed in a third of patients with EAC. Therapeutic trials are already underway using GPA33-directed therapies in other cancers and our finding of EAC-specific expression of this cell surface protein in a significant proportion of patients provides evidence to extend trials to consider patients with EAC.

We provide the first combined global analysis of protein and RNA expression in EAC and identify several novel genes with high or low protein-to-RNA expression ratios suggesting the decorrelation of RNA and protein abundance. We further confirm these findings are specific to EAC for several candidate genes by undertaking a combined analysis of protein and RNA expression across multiple normal tissue types using external datasets and confirm these genes are not dysregulated by somatic mutation. We finally identify AGMAT as over-expressed at the protein level in EAC compared to adjacent normal tissues and identify the mechanism of over-expression as post-transcriptional and due to its EAC-specificity may offer a targetable vulnerability.

## Data Availability

All data produced in the present study are available upon reasonable request to the authors.

## List of abbreviations

EAC: Esophageal adenocarcinoma
TMT: Tandem Mass Tag
RT: Room temperature
MS: Mass spectrometry
AGC: Automated gain control
FDR: False discovery rate
TvE: Tumor vs normal esophagus
TvG: Tumor vs normal gastric tissue
TMM: Trimmed-median of M-values
IHC: Immunohistochemistry
TMAs: Tissue microarrays
TPM: Transcript per million
GTEx consortium: Genotype-Expression consortium dataset
ICGC ESAD: International Cancer Genome Consortium Esophageal Adenocarcinoma cancer genome project
hg38: Human reference genome version 38
GATK: Genome Analysis Toolkit
ITLN1: Intelectin-1
VIL1: Villin
RBM: RNA binding motif
Insulin-Like Growth Factor-Binding Protein 1: IGF2BP1
GPA33: Glycoprotein A33

## Declarations

### Ethics approval and consent to participate

Institutional Review Board Approval. All patients gave prospective written informed consent to the use of tissue, clinical data, and publication with ethical and research governance approvals in place from the Lothian Local Research Ethics Committee (UK, REC references 06/S1101/16), Tayside Committee on Medical Research Ethics (UK, REC 10/S1402/33), Cambridgeshire 4 Research Ethics Committee (UK, REC 10/H0305/1) and the NHS Lothian Research and Development Office (UK, R&D ID 2006/W/PA/01, R&D ID 2011/W/ON.27).

### Consent for publication

Patients were de-identified at the time of consent to the use of tissue and clinical data and therefore no patient-identifiable data have been included. Consent to publication of results arising from the use of tissue and data were prospectively obtained.

### Availability of data and materials

All data produced in the present study are available upon reasonable request to the authors.

### Authors’ contributions

M.Y.M, R.O and J. A. conceived of and initiated the project. J. A. and R.O. coordinated and supervised the project. R.O., G.B., M.Y.M, MK recruited patients, collected the tissue, collated the online studies, developed the computational approach, processed the data, and created the figures. Raw RNAseq and DNAseq data were generated by the OCCAMS consortium. G.M., J.F., L.H., L.U., M.G-H, contributed to both sample processing, generating and processing the raw MS data. V.S., M.A. and S.A.S. performed immunohistochemistry and provided expert pathology review. B.V. and T.R.H obtained funding for the project. The funding bodies were not involved in the design of the study, analysis, interpretation of data, and writing this manuscript. The manuscript was drafted by M. Y. M., R.O., J. A, G.B., S.A.S, T.H. and R.C.F (OCCAMS). The final manuscript draft was reviewed and approved by all authors.

The Oesophageal Cancer Clinical and Molecular Stratification (OCCAMS) Consortium is composed by:

Rebecca C. Fitzgerald^1^, Paul A.W. Edwards^1,2^, Nicola Grehan^1^, Barbara Nutzinger^1^, Elwira Fidziukiewicz^1^, Aisling M Redmond^1^, Sujath Abbas^1^, Adam Freeman^1^ Elizabeth C. Smyth^5^, Maria O’Donovan^1,3^, Ahmad Miremadi^1,3^, Shalini Malhotra^1,3^, Monika Tripathi^1,3^, Aisling Redmond^1^, Calvin Cheah^1^, Hannah Coles^1^ Connor Flint^1^ Matthew Eldridge^2^, Maria Secrier^2^, Ginny Devonshire^2^, Sriganesh Jammula^2^, Jim Davies^4^, Charles Crichton^4^, Nick Carroll^5^, Richard H.Hardwick^5^, Peter Safranek^5^, Andrew Hindmarsh^5^, Vijayendran Sujendran^5^, John Bennett^5^, Stephen J. Hayes^6,13^, Yeng Ang^6,7,26^, Andrew Sharrocks^26^, Shaun R. Preston^8^, Izhar Bagwan^8^, Vicki Save^9^, Richard J.E. Skipworth^9^, Ted R. Hupp^20^, J. Robert O’Neill^5,9,20^, Olga Tucker^10, 29^, Andrew Beggs^10, 25^, Philippe Taniere^10^, Sonia Puig^10^, Gianmarco Contino^10^, Timothy J. Underwood^11, 12^, Robert C. Walker^11, 12^, Ben L. Grace^11^, Jesper Lagergren^14,22^, James Gossage^14,21^, Andrew Davies^14,21^, Fuju Chang^14,21^, Ula Mahadeva^14^, Vicky Goh^21^, Francesca D. Ciccarelli^21^, Grant Sanders^15^, Richard Berrisford^15^, David Chan^15^, Ed Cheong^16^, Bhaskar Kumar^16^, L. Sreedharan^16^ Simon L Parsons^17^, Irshad Soomro^17^, Philip Kaye^17^, John Saunders^6, 17^, Laurence Lovat^18^, Rehan Haidry^18^, Michael Scott^19^, Sharmila Sothi^23^, Suzy Lishman^2^, George B. Hanna^27^, Christopher J. Peters^27^,Krishna Moorthy^27^, Anna Grabowska^28^, Richard Turkington^30^, Damian McManus^30^, Helen Coleman^30^, Russell D Petty^32^, Freddie Bartlett^33^

^1^ Medical Research Council Cancer Unit, Hutchison/Medical Research Council Research Centre, University of Cambridge, Cambridge, UK

^2^ Cancer Research UK Cambridge Institute, University of Cambridge, Cambridge, UK

^3^ Department of Histopathology, Addenbrooke’s Hospital, Cambridge, UK

^4^Department of Computer Science, University of Oxford, UK, OX1 3QD

^5^Cambridge University Hospitals NHS Foundation Trust, Cambridge, UK, CB2 0QQ

^6^Salford Royal NHS Foundation Trust, Salford, UK, M6 8HD

^7^Wigan and Leigh NHS Foundation Trust, Wigan, Manchester, UK, WN1 2NN

^8^Royal Surrey County Hospital NHS Foundation Trust, Guildford, UK, GU2 7XX

^9^Edinburgh Royal Infirmary, Edinburgh, UK, EH16 4SA

^10^University Hospitals Birmingham NHS Foundation Trust, Birmingham, UK, B15 2GW

^11^University Hospital Southampton NHS Foundation Trust, Southampton, UK, SO16 6YD

^12^Cancer Sciences Division, University of Southampton, Southampton, UK, SO17 1BJ

^13^Faculty of Medical and Human Sciences, University of Manchester, UK, M13 9PL

^14^ Guy’s and St Thomas’s NHS Foundation Trust, London, UK, SE1 7EH

^15^Plymouth Hospitals NHS Trust, Plymouth, UK, PL6 8DH

^16^Norfolk and Norwich University Hospital NHS Foundation Trust, Norwich, UK, NR4 7UY

^17^Nottingham University Hospitals NHS Trust, Nottingham, UK, NG7 2UH

^18^University College London, London, UK, WC1E 6BT

^19^Wythenshawe Hospital, Manchester, UK, M23 9LT

^20^Edinburgh University, Edinburgh, UK, EH8 9YL

^21^King’s College London, London, UK, WC2R 2LS

^22^Karolinska Institute, Stockholm, Sweden, SE-171 77

^23^University Hospitals Coventry and Warwickshire NHS, Trust, Coventry, UK, CV2 2DX

^24^Peterborough Hospitals NHS Trust, Peterborough City Hospital, Peterborough, UK, PE3 9GZ

^25^Institute of Cancer and Genomic sciences, University of Birmingham, B15 2TT

^26^GI science centre, University of Manchester, UK, M13 9PL.

^27^Department of Surgery and Cancer, Imperial College, London, UK, W2 1NY

^28^Queen’s Medical Centre, University of Nottingham, Nottingham, UK

^29^Heart of England NHS Foundation Trust, Birmingham, UK, B9 5SS.

^30^Centre for Cancer Research and Cell Biology, Queen’s University Belfast, Northern Ireland BT7 1NN.

^32^Tayside Cancer Centre, Ninewells Hospital and Medical School, Dundee, DD1 9SY

^33^ Portsmouth Hospitals NHS Trust, Portsmouth, PO6 3LY

## Acknowledgments

Additional infrastructure support and assistance with tissue collection, storage, and tissue microarray construction were provided from the Cancer Research UK–funded Experimental Cancer Medicine Centers in Edinburgh and Cambridge. The authors would like to thank the PL-Grid Infrastructure and CI-TASK, Poland for providing their hardware and software resources and the Human Research Tissue Bank from Addenbrookes Hospital which is supported by the UK National Institute for Health Research (NIHR) Cambridge Biomedical Research Center.

The Genotype-Tissue Expression (GTEx) Project was supported by the Common Fund of the Office of the Director of the National Institutes of Health, and by NCI, NHGRI, NHLBI, NIDA, NIMH, and NINDS.

## Supplementary legends

**Supplementary table 1:** Relative tissue expression ratios of all quantified proteins.

**Supplementary figure 1:**
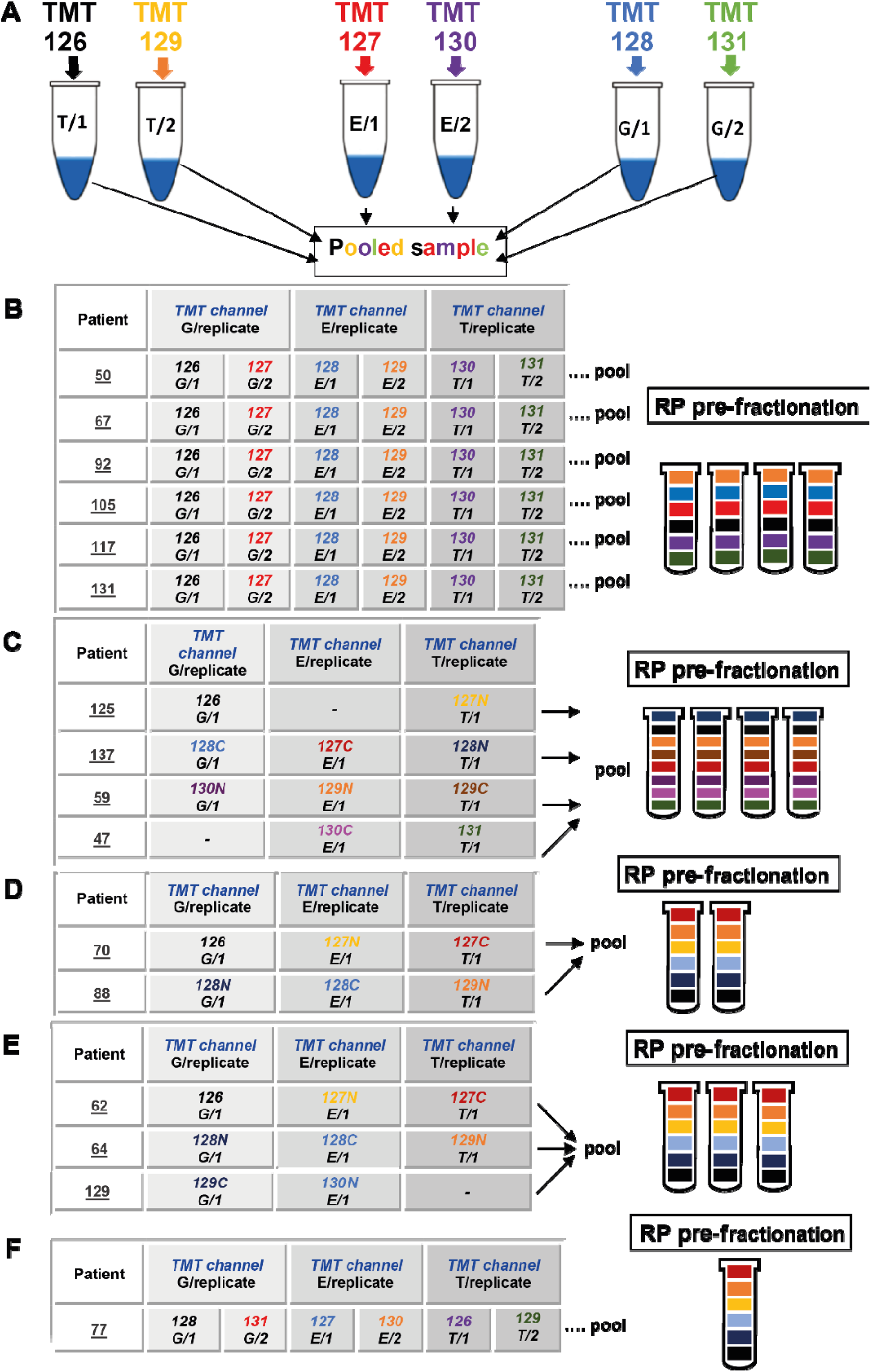
Individual patient TMT labeling and fractionation protocol for EAC, normal esophageal, and gastric samples. **(a)** Labeling strategy from initial 7 patients from prior published study^13^. **(b-e)** Labeled sample pools were each separated by reverse phase (RP) basic pH fractionation and then each fraction was subjected to LC-MS/MS followed by either a six-plex strategy **(b)** or ten-plex strategy with empty channels **(c-e)**. T-EAC tissue, E - normal Esophagus, G - normal stomach.

**Supplementary figure 2:**
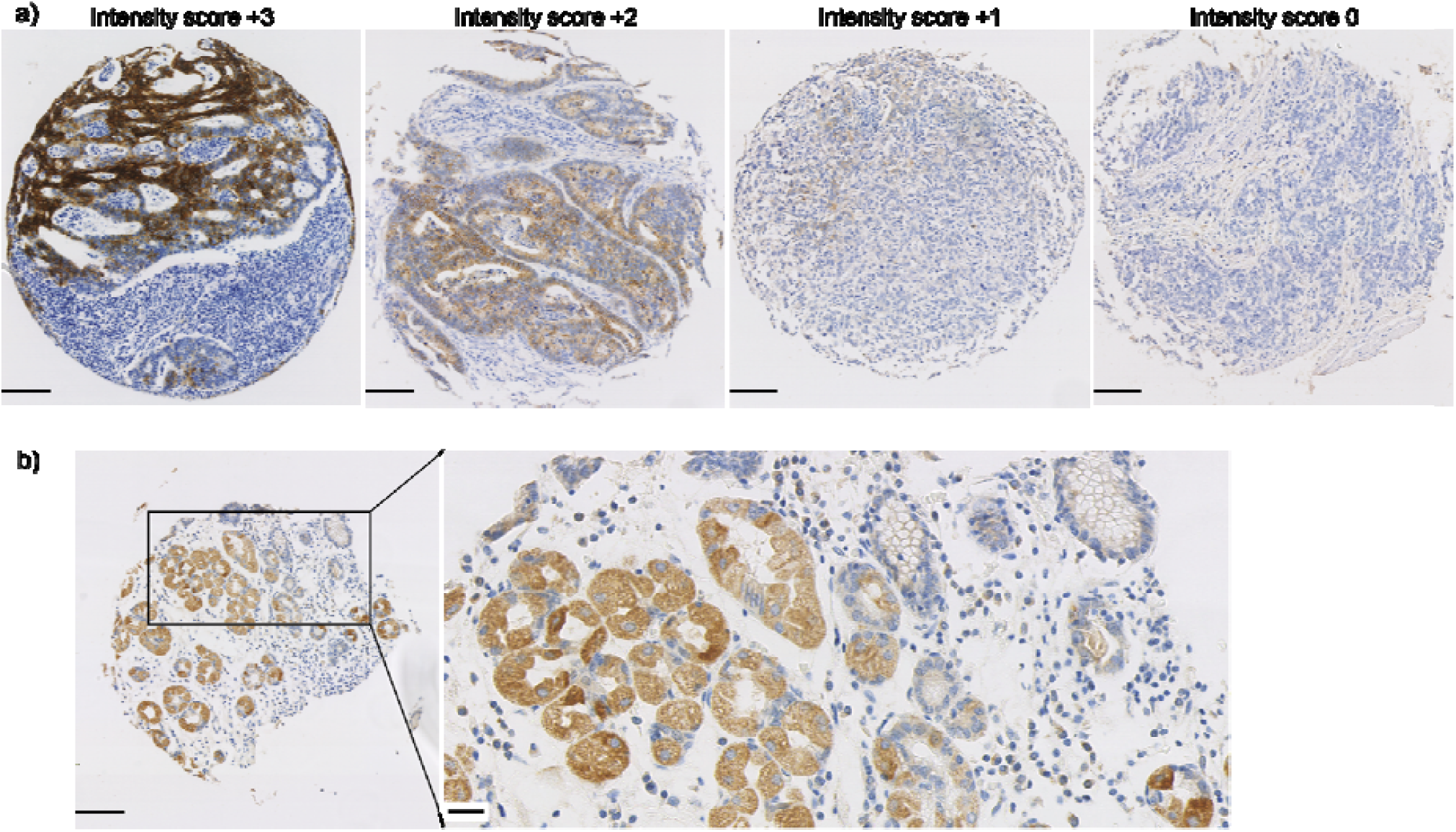
Representative TMA Immunohistochemistry images. **(a)** Core images showing examples of intensity evaluation used to score the samples, +3 and +2: high, +1: low, and 0: negative. **(b)** Normal gastric glands showing high expression of IGF2BP1).

**Supplementary figure 3:**
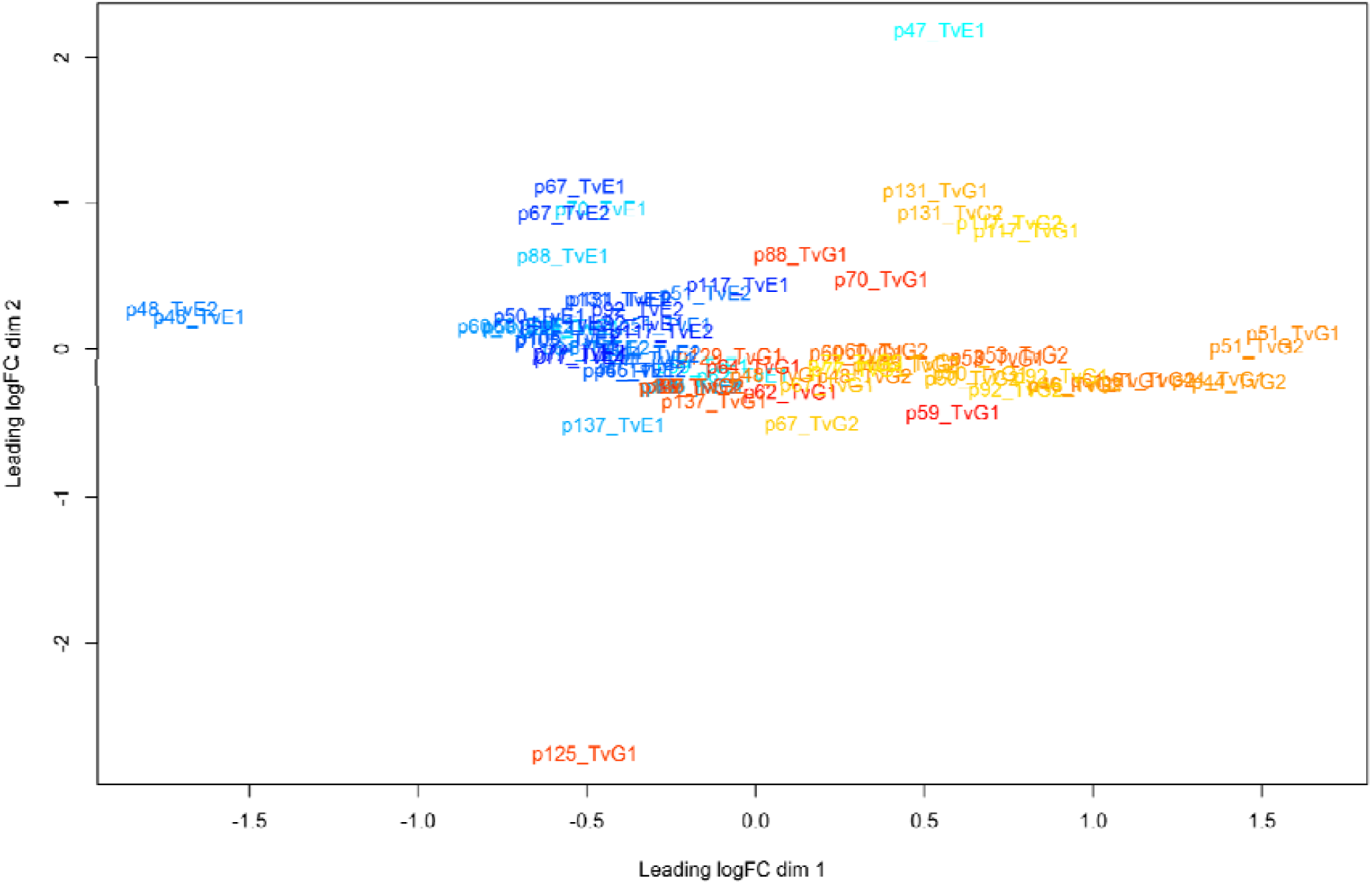
Multi-dimensional scaling (MDS) plot of individual patient technical replicate quantitative ratios. TvE - EAC vs. normal esophagus, TvG - EAC vs. normal stomach, p - patient number.

**Supplementary figure 4:**
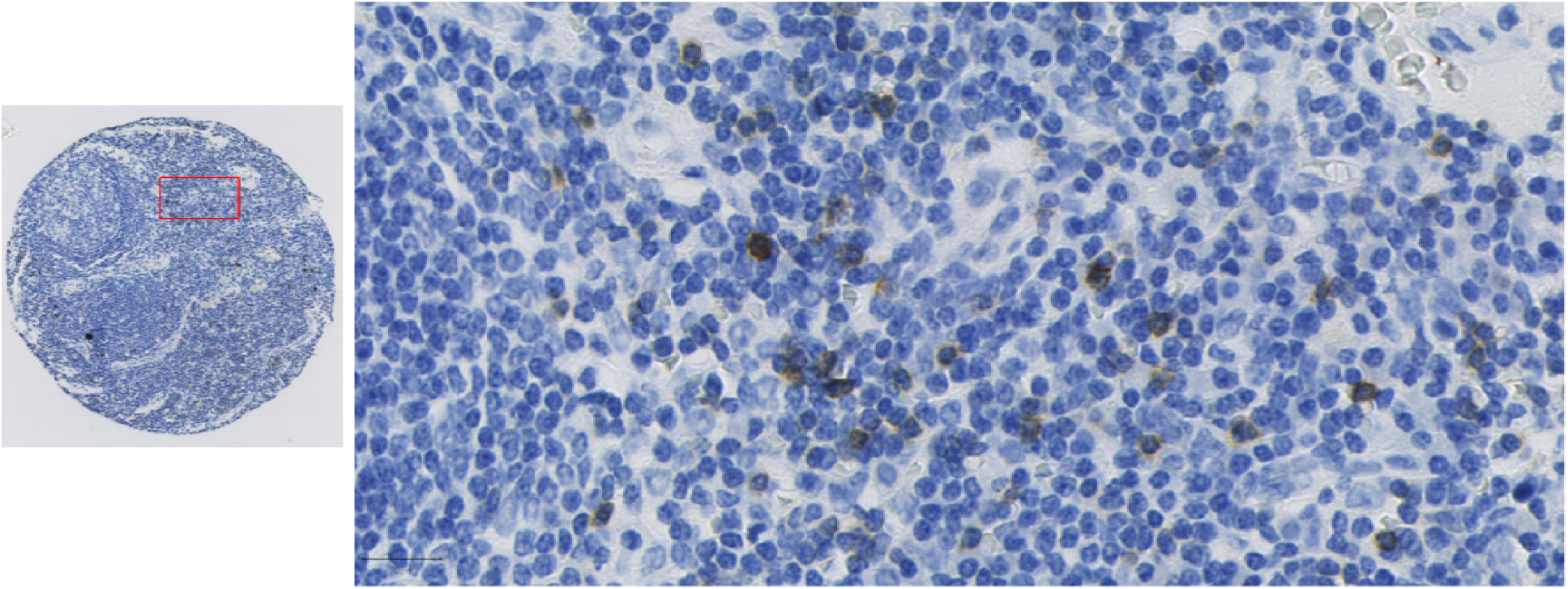
GPA33 staining in uninvolved lymph nodes. Representative image of GPA33 staining arising in lymphocytes.

**Supplementary figure 5:**
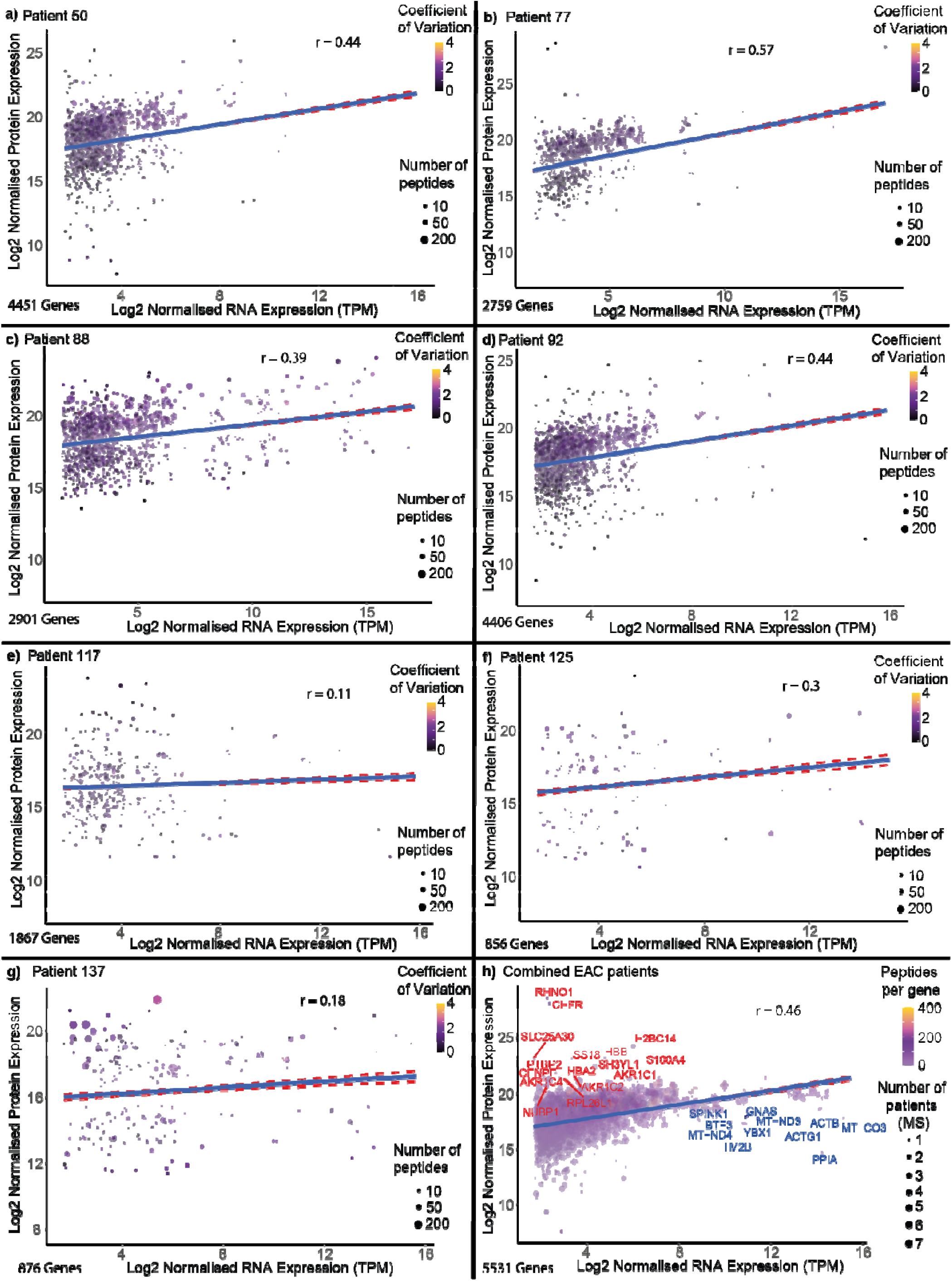
Correlation of Protein and RNA abundance in EAC. Patient-matched data are displayed in panels (a) - (g). Each point represents a gene and points have been sized according to the number of peptides quantified and colored according to the relative coefficient of variation across peptide quantities. (h) - Combined data from 7 patients. Each point has been sized according to the number of patients both Protein and RNA were quantified in and colored according to the number of peptides quantified.

**Supplementary figure 6:**
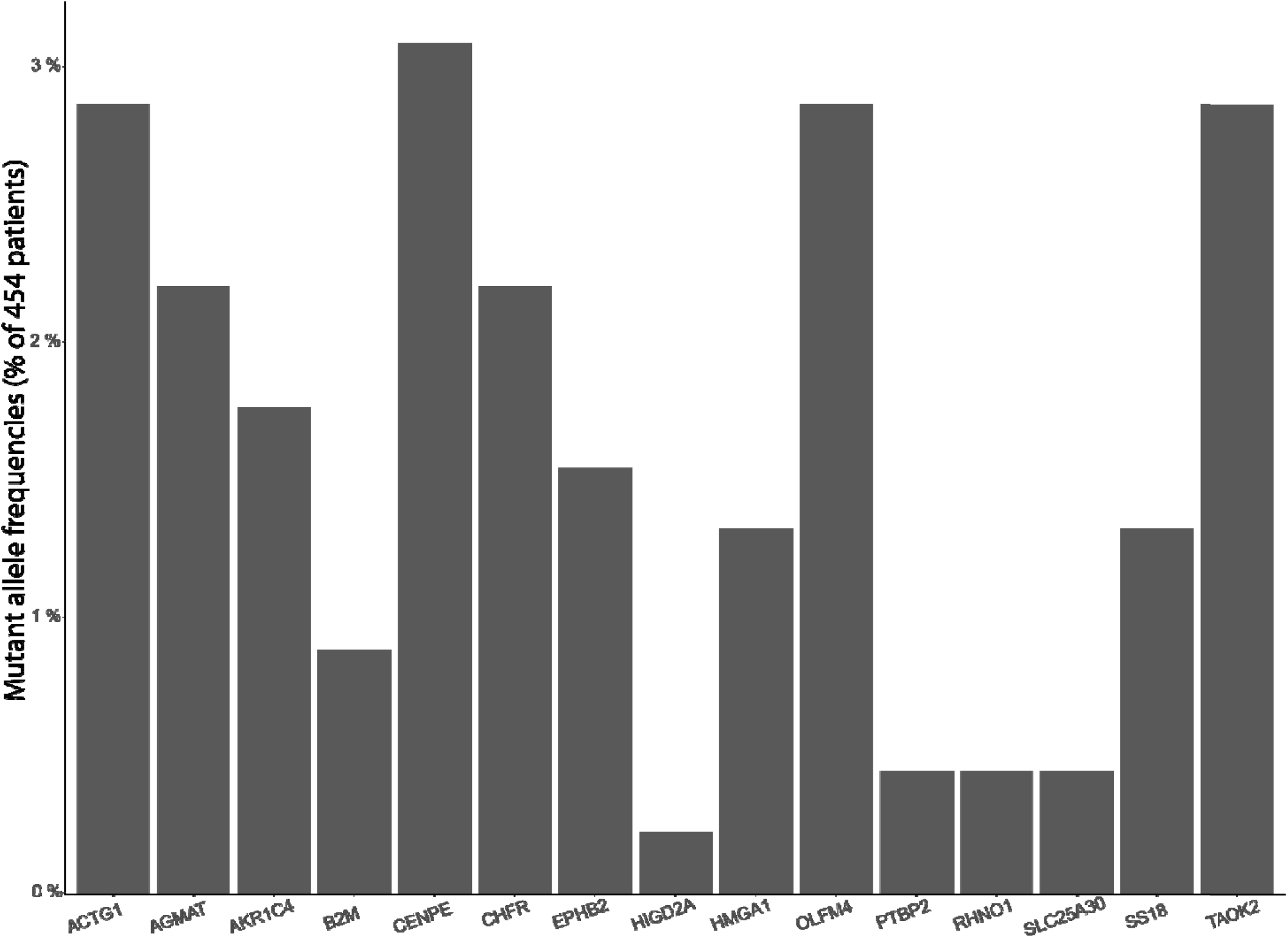
Somatic mutations in outlier genes. The percentage of EAC patients affected by a mutation for all outlier genes was calculated using whole-genome sequencing from a cohort of 454 patients.

