## Supplemental Figure 1 for "Multi-omics integration in Esophageal Adenocarcinoma reveals therapeutic targets and EAC-specific regulation of protein abundances"

A

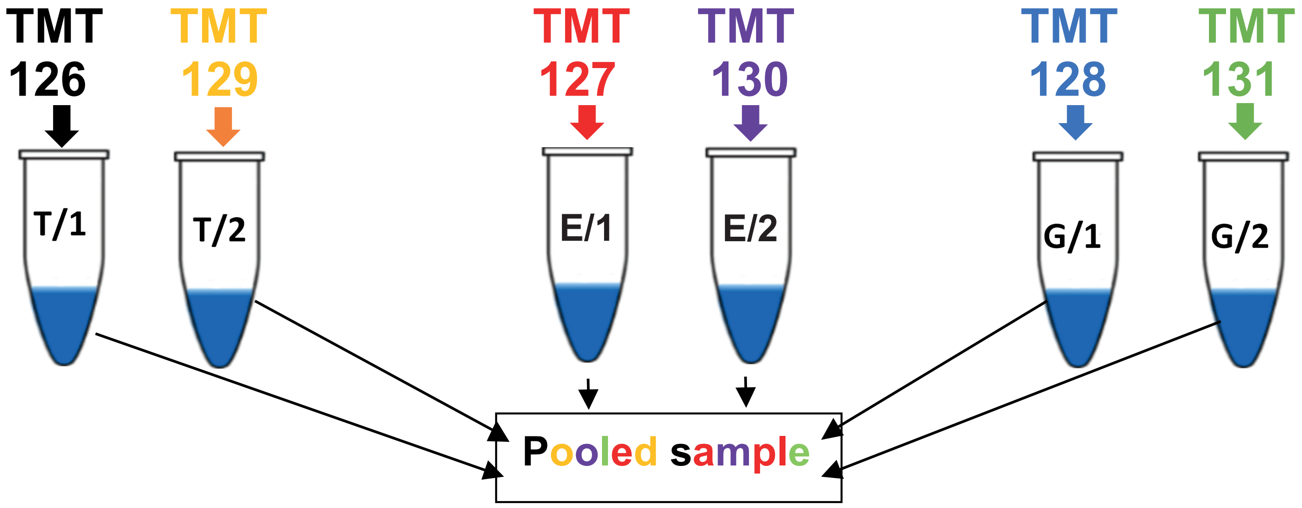


B

| Patient | TMT channel G/replicate | | TMT channel E/replicate | | TMT channel T/replicate | | |
|---|---|---|---|---|---|---|---|
| 50 | 126 G/1 | 127 G/2 | 128 E/1 | 129 E/2 | 130 T/1 | 131 T/2 | .... pool |
| 67 | 126 G/1 | 127 G/2 | 128 E/1 | 129 E/2 | 130 T/1 | 131 T/2 | .... pool |
| 92 | 126 G/1 | 127 G/2 | 128 E/1 | 129 E/2 | 130 T/1 | 131 T/2 | .... pool |
| 105 | 126 G/1 | 127 G/2 | 128 E/1 | 129 E/2 | 130 T/1 | 131 T/2 | .... pool |
| 117 | 126 G/1 | 127 G/2 | 128 E/1 | 129 E/2 | 130 T/1 | 131 T/2 | .... pool |
| 131 | 126 G/1 | 127 G/2 | 128 E/1 | 129 E/2 | 130 T/1 | 131 T/2 | .... pool |

RP pre-fractionation

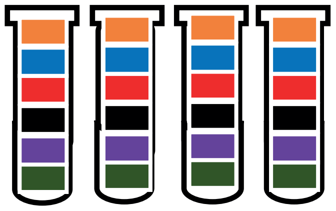

C

| Patient | TMT channel G/replicate | TMT channel E/replicate | TMT channel T/replicate |
|---|---|---|---|
| 125 | 126 G/1 | - | 127N T/1 |
| 137 | 128C G/1 | 127C E/1 | 128N T/1 |
| 59 | 130N G/1 | 129N E/1 | 129C T/1 |
| 47 | - | 130C E/1 | 131 T/1 |

→ pool

RP pre-fractionation

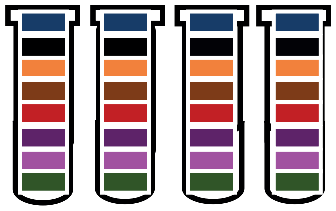

D

| Patient | TMT channel G/replicate | TMT channel E/replicate | TMT channel T/replicate |
|---|---|---|---|
| 70 | 126 G/1 | 127N E/1 | 127C T/1 |
| 88 | 128N G/1 | 128C E/1 | 129N T/1 |

→ pool

RP pre-fractionation

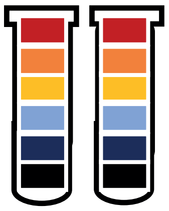

E

| Patient | TMT channel G/replicate | TMT channel E/replicate | TMT channel T/replicate |
|---|---|---|---|
| 62 | 126 G/1 | 127N E/1 | 127C T/1 |
| 64 | 128N G/1 | 128C E/1 | 129N T/1 |
| 129 | 129C G/1 | 130N E/1 | - |

→ pool

RP pre-fractionation

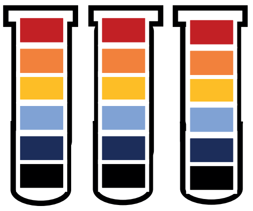

F

| Patient | TMT channel G/replicate | | TMT channel E/replicate | | TMT channel T/replicate | | |
|---|---|---|---|---|---|---|---|
| 77 | 128 G/1 | 131 G/2 | 127 E/1 | 130 E/2 | 126 T/1 | 129 T/2 | .... pool |

RP pre-fractionation

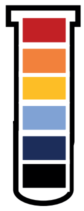
