## Supplementary figures and images for "Multi-omics integration in Esophageal Adenocarcinoma reveals therapeutic targets and EAC-specific regulation of protein abundances"

### Supplemental Figure 2

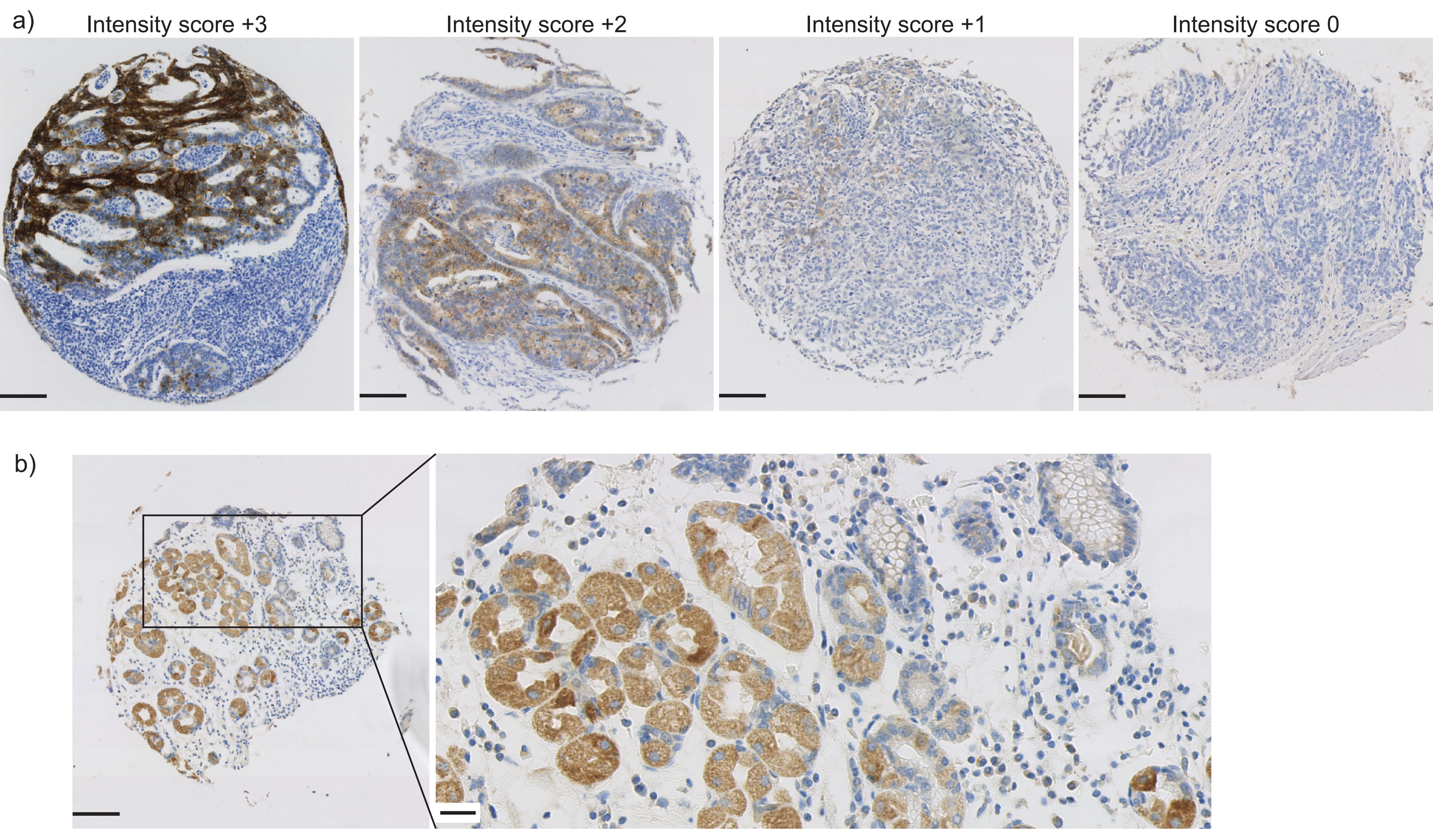

### Supplemental Figure 3

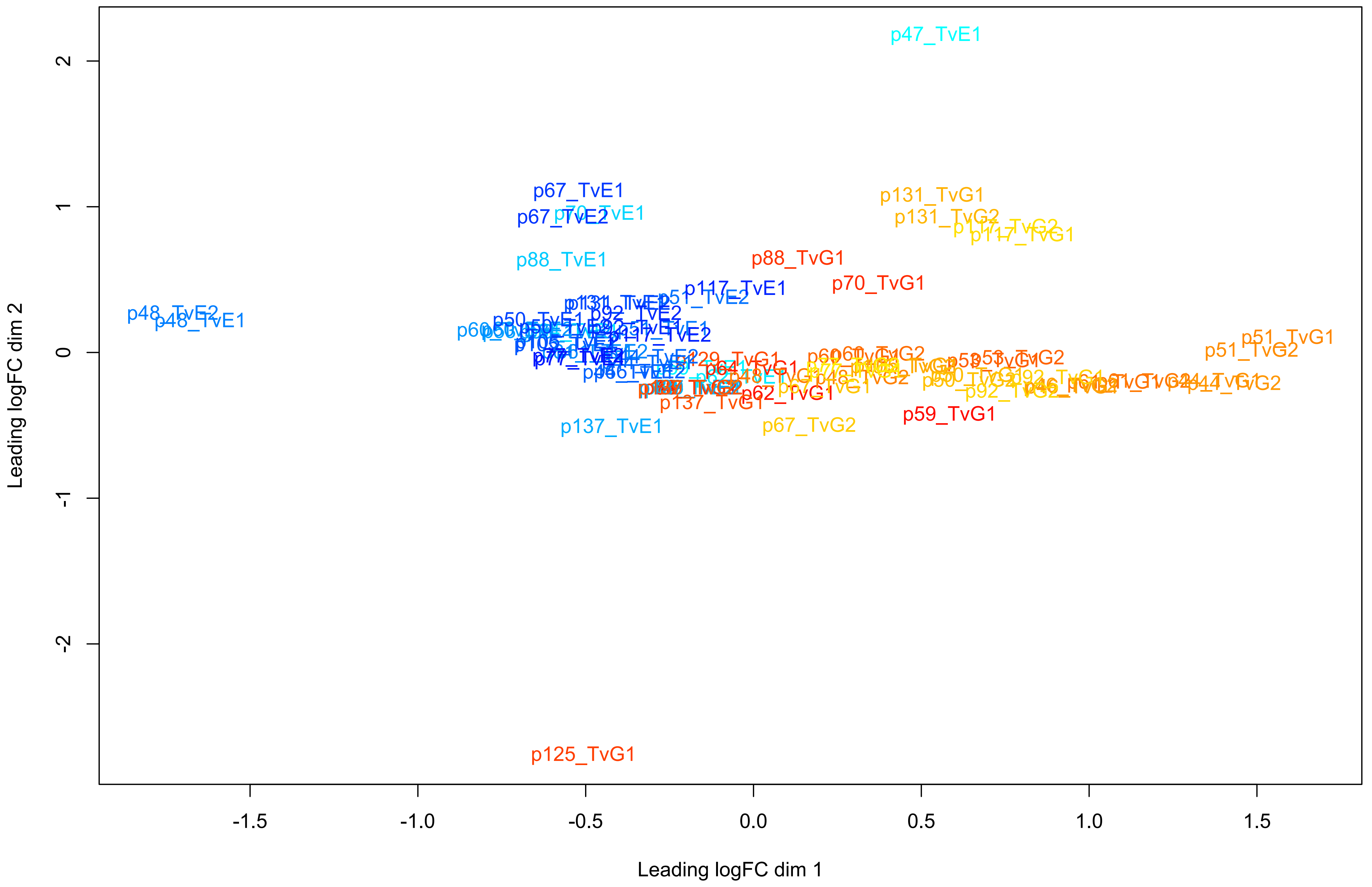

### Supplemental Figure 4

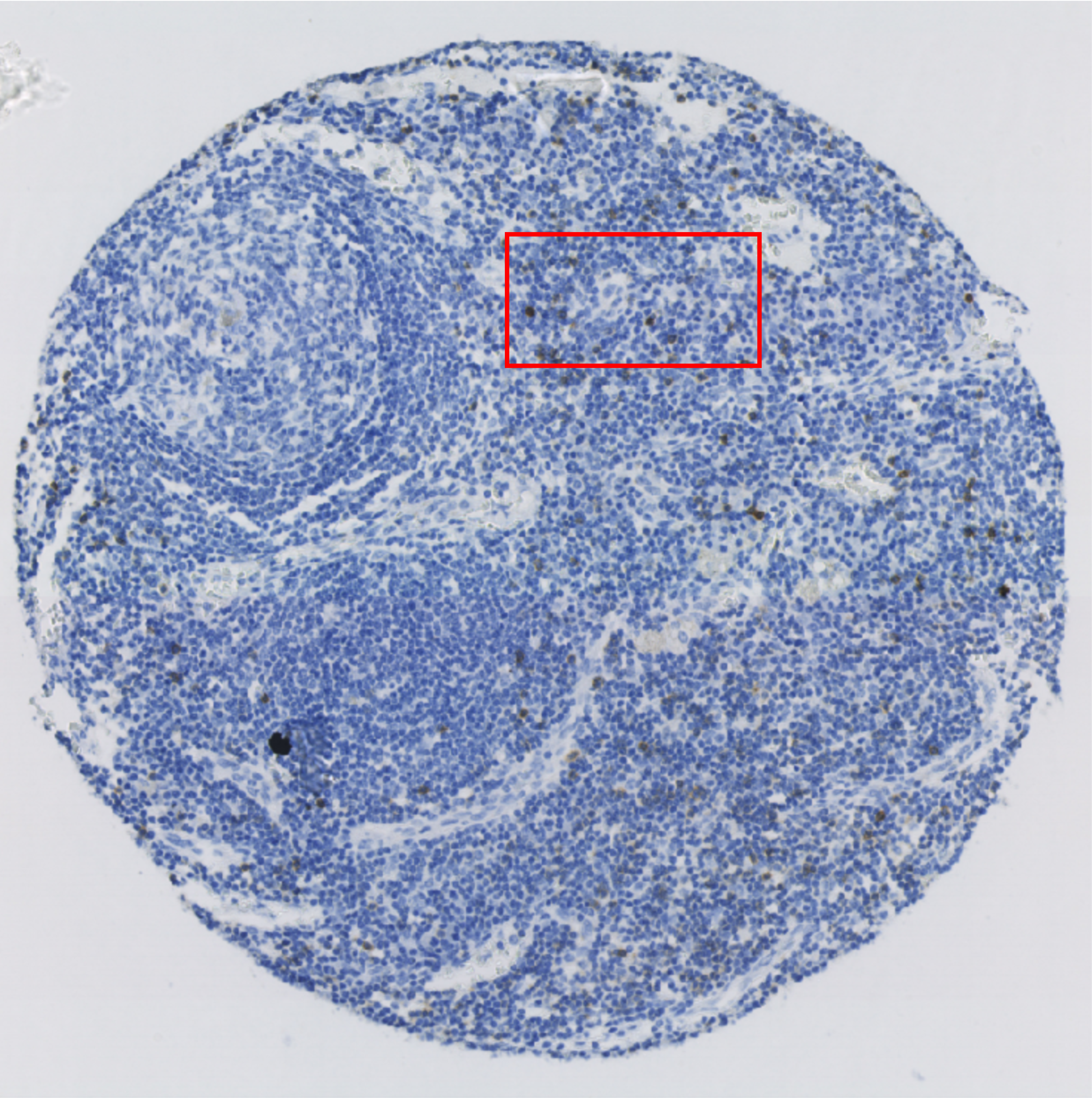

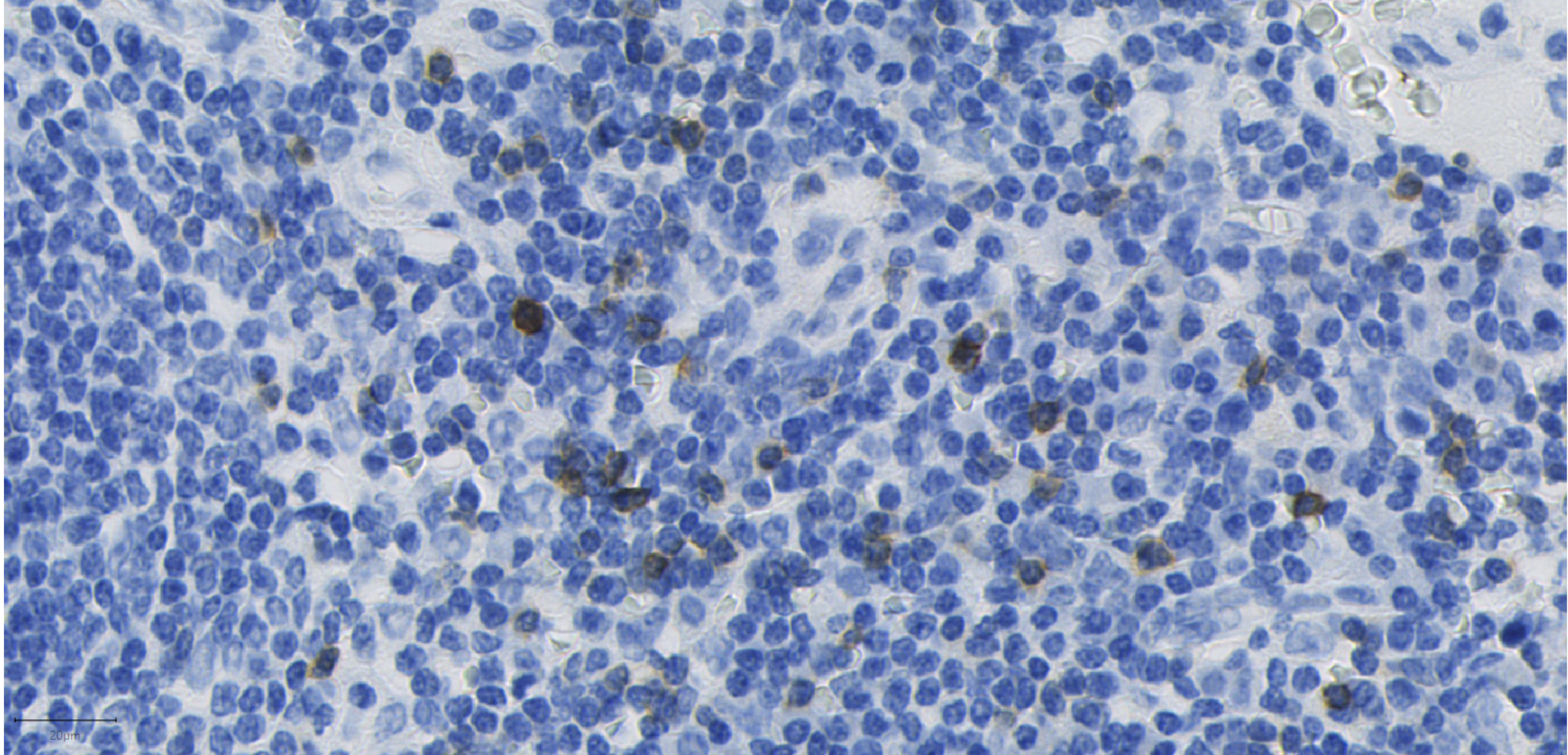

### Supplemental Figure 5

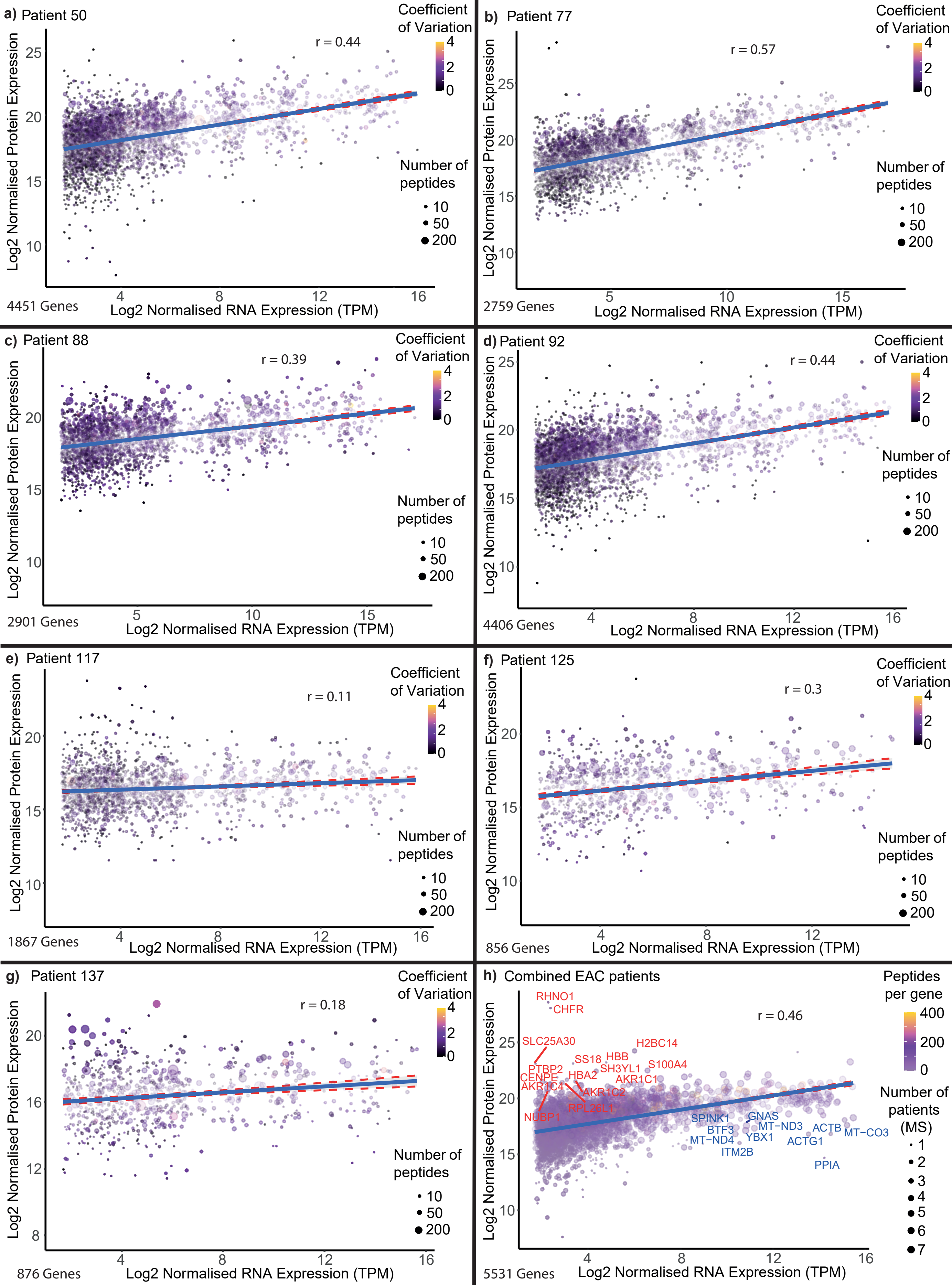

### Supplemental Figure 6

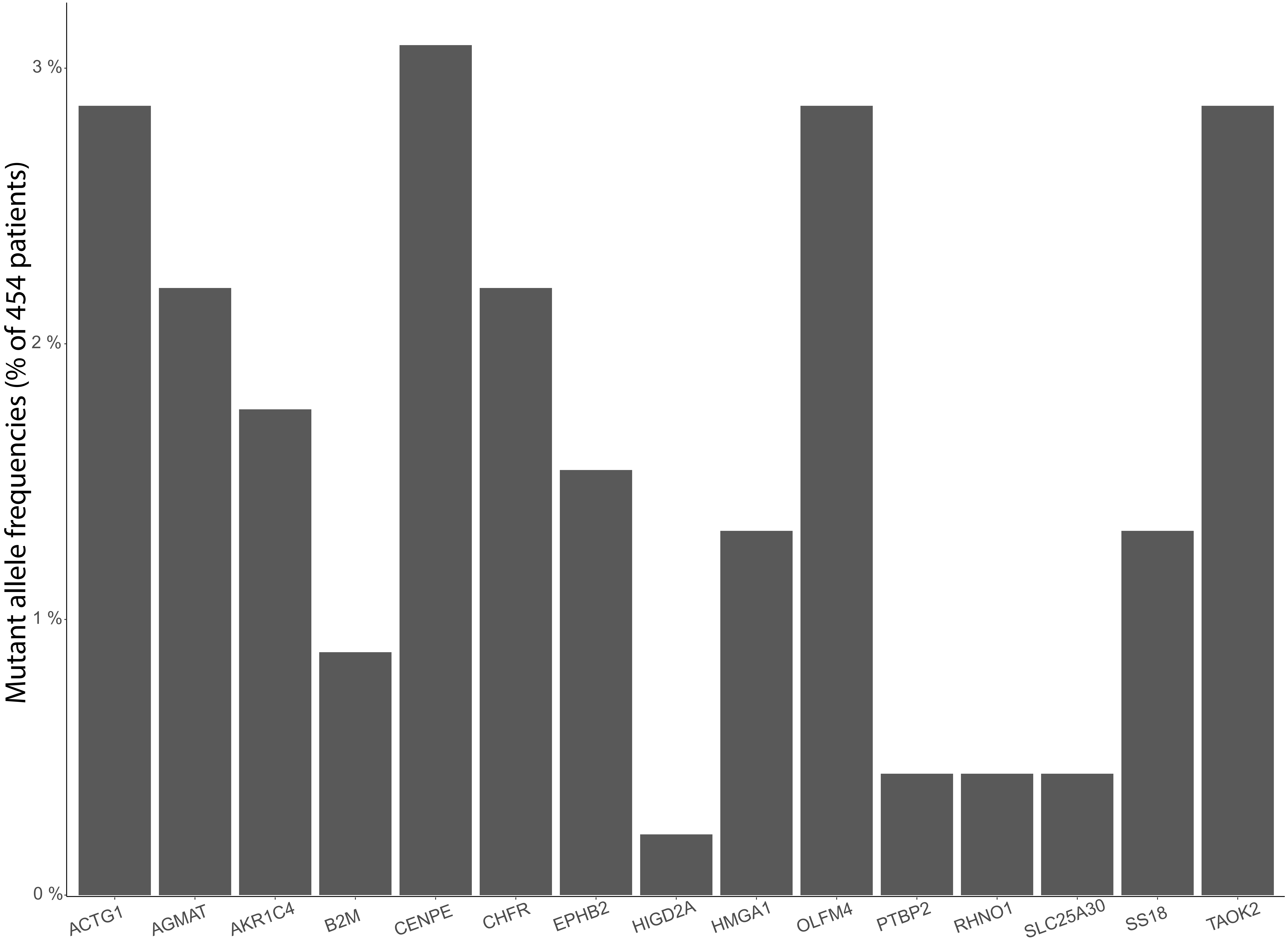
