## Supplemental Methods for "Multi-omics integration in Esophageal Adenocarcinoma reveals therapeutic targets and EAC-specific regulation of protein abundances"

### Supplementary methods

**Protocol for modified Filter-aided sample preparation for Oesophageal and Gastric Tissues for Mass Spectrometry.**

**Tissue Lysis**

1) Cut tissue aliquots if required on dry ice (aim ~20mg). Place directly into Matrix D homogenization tubes. Using one homogenization tube as a blank, weigh the tissue aliquots.
2) Homogenise using the MP Bio Homogeniser (Irvine, CA, USA) on “small intestine settings” – 6 ms-1 for 40 seconds at room temperature. Check to ensure the tissue has been broken down. Repeat once or twice more as necessary.
3) Add 20:1 (volume µL : mass mg) complete lysis buffer: tissue using filter tips and pipette to dissolve the tissue e.g. 600 µL:30mg.
4) Gently mix the homogenate in the Matrix D tubes for ~20 minutes at room temperature on a rotating mixer. Inspect to make sure the tissue is well dissolved.
5) With the tubes resting in ice, sonicate using a needle sonicator (Bioruptor ,Diagenode, Denville, NJ, USA) with the settings amplitude 10.0, x1, 20-30s).
6) Heat the lysate at 95 degrees C for 5 mins.
7) Aspirate the supernatant – transfer it to a labeled Eppendorf tube. Centrifuge for 5 mins at 14,000 x g (200C). The pellet should be very small. If large then this should be resuspended in a small volume of lysis buffer, sonicated, and recentrifuged. (Pool all the lysate from one tissue biopsy together).

**Buffer Exchange**

8) Transfer 30µL of clarified lysate to each labeled 30kDa NMWC filter (Millipore, Watford, UK) required. Aim for 6 aliquots for MS (finish at tryptic peptides), 5 for reverse-phase protein array or Western (stop at UL stage), and 1 for RC-DC (stop once proteins pooled pre-trypsinization). Residual SDS-lysate can be snap-frozen in reserve.
9) Add 200µL of Urea Solution A (UA) and centrifuge for 15 minutes, 14,000g (200C). Repeat x1.
10) The 30µL of remaining lysate is the buffer exchanged protein solution.
11) Discard the flow-through from the collection tube.
12) Add 100μl IAA solution and place in the polystyrene tube holder. Protect from light and mix on the orbital shaker at maximum for 1 min and incubate without mixing for 20 min at room temperature.
13) Centrifuge the filter units at 14,000 x g for 10 min.
14) Add 100μl of UA to the filter unit and centrifuge at 14,000 x g for 15 min. Repeat x 2.
15) Add 100μl of TEAB solution (50mM TEAB in dH2O) to the filter unit and centrifuge at 14,000 x g for 10 min. Repeat x2.
16) Aspirate the residue and pool together in an Eppendorf-brand microcentrifuge tube. Vortex and then quantify using 12.5uL of lysate and the RC-DC method (equivalent to a 1 in 2 dilutions).
17) Proceed to the trypsinization stage using known quantities of protein.

**Trypsinization**

18) Resuspend the lyophilized trypsin (20 µg per glass vial) in 10µL of 1M HCL (MS grade stock). Vortex. Dilute down to 1ml total volume using TRB.
19) Add Trypsin solution to Eppendorf containing a known quantity of protein lysate in a 100 protein : 1 trypsin ratio (w/w).
20) Final composition Trypsin at 1% of protein mass, 20-50mM TEAB depending on ratio of TRB to lysate, ~3-9% Acetonitrile, sample protein.
21) Check the pH of the trypsin/protein solution by touching the pH paper with the pipette (expect pH~8).
22) Vortex the labelled trypsin tubes and place each closed tube in a rack in an incubator at 37 degrees C for 4-18 hours.
23) Prepare for peptide desalting (or Snap freeze for desalting elsewhere).
24) (Optional to reduce the TEAB). Evaporate the tubes to dryness using a vacuum centrifuge (SpeedVac, ThermoFisher Scientific, Loughborough, UK) on “medium” heat setting. Resuspend in 200uL of MS grade H2O and centrifuge to dryness again. Repeat x2. Snap freeze the dried tryptic peptides.

**HPLC conditions and mobile phases.**

The nano chromatographic separation was performed using a nano-RSLC UltiMate system (ThermoFisher Scientific, Loughborough, UK). For sample loading and desalting, a PepMap C18 Trap-column (ThermoFisher Scientific, Loughborough, UK) of 300 µm ID x 5 mm length, 5 µm particle size, and 100 Å pore size were used. Nano HPLC Separation of labeled and digested proteins was performed using a C18 µPAC separation column (PharmaFluidics, Ghent, Belgium). The dimensions of the used µPAC column were: 5 µm pillar diameter, 2.5 µm inter-pillar distance, 18 µm pillar height; 315 µm bed channel width, and 200 cm column length. The pillars are superficially porous and end-capped with C18 chains.

The aqueous loading mobile phase contained 2% ACN,0.1% TFA, and 0.01% HFBA cooled to 3°C, as described earlier^1^. The mobile phase transferred the sample from the sample loop to the trap column by the loading pump at 30 µl/min to the trapping column, which was operated in the column oven at 50°C. The trapping time was set to 5 min, which was sufficient to load the sample to the trapping column and wash possible salts and contaminants. The cooled mobile phase enabled the trapping of the hydrophilic analytes at enhanced oven temperature. Peptides were separated and analyzed using positive nano HPLC-ESI-MS. The gradient elution was performed by mixing two mobile phases composed of the following solvents:

- Mobile phase A (MPA): 95% H2O, 5% ACN, 0.1% FA;
- Mobile phase B (MPB): 95% ACN, 5% MeOH, 0.1% FA.

At 240 minutes, the trapping column was switched back into the separation flow and equilibrated for the following run.

The nano HPLC separation was performed on the µPAC separation column with a 2 m separation path. Due to the long separation path, the void volume of these columns is higher in comparison to conventionally packed nano separation columns. Therefore, the flow rate of the nano HPLC pump was set to 800 nl/min for the first 10 minutes with a gradual lowering to 600 nl/min for separation purposes. The gradual increase of the flow rate at the end of the separation run to 800 nl/min is performed for speeding up the equilibration of the separation column. Details on developing and testing different gradients for the 200-cm µPAC column are described by Tóth *et al.*^2^

Detection of eluting peptides is performed using both UV at 214 nm (3nl UV cell, ThermoFisher Scientific, Germering, Germany) and mass spectrometry (Q-Exactive Plus, ThermoFisher Scientific, Bremen, Germany). The separation gradient applied is described in **Supplementary Table 1**.

| **Time (min)** | **Flow (nl/min)** | **%A** | **%B** |
| --- | --- | --- | --- |
| 0 | 800 | 97 | 3 |
| 10 | 800 | 97 | 3 |
| 12 | 600 | 97 | 3 |
| 100 | 600 | 90 | 10 |
| 180 | 600 | 70 | 30 |
| 205 | 600 | 20 | 80 |
| 210 | 800 | 10 | 90 |
| 219 | 800 | 10 | 90 |
| 240 | 800 | 97 | 3 |

**Supplementary Table 1.** Detailed separation gradient used in the Q-Exactive mass spectrometry.
